# Exon Proteogenomic Abundance Predicts Advanced Heart Failure in Titin Cardiomyopathy

**DOI:** 10.1101/2025.11.02.25338804

**Authors:** Peter Deckerman, Jihyeon Lee, Aleksandra Binek, Sarah Parker, Daniel J. Luthringer, Michelle M. Kittleson, Donald Richards, Ian Williamson, Daria Ma, Lawrence S.C. Czer, Nan Jiang, Jason Cowan, Carl A. Starkey, Ray E. Hershberger, Jennifer E. Van Eyk, Evan P. Kransdorf

**Affiliations:** Smidt Heart Institute, Cedars-Sinai Medical Center, Los Angeles, CA 90048, USA; Davis Heart and Lung Research Institute, The Ohio State University, Columbus, OH 43210, USA

## Abstract

**Background and Aims:** Titin (TTN) truncating variants (TTN-TV) are the leading genetic cause of dilated cardiomyopathy (DCM) and confer significant risk for advanced heart failure (AHF). TTN-TV penetrance links to the expression level of the affected exon, quantified by “percent spliced in” (PSI). We hypothesized that recalculating PSI via long-read RNA sequencing and including all 15 TTN isoforms would improve predictions of DCM penetrance and AHF expressivity. Given potential for discordance between transcript and protein abundance, we also compared PSI to exon-specific TTN peptide abundance and evaluated a hybrid proteogenomic metric.

**Methods:** We performed long-read RNA sequencing (9 organ donor controls, 25 DCM explants) and mass spectrometry-based peptide mapping (14 controls, 14 DCM explants) on human cardiac tissue to identify and quantify TTN isoforms. Exon abundance was quantified using a novel long-read RNA metric incorporating all 15 TTN isoforms (PSI-DCM-LR-15) and quantile peptide intensity (QPI), which reflects exon-specific peptide abundance. These were integrated into a hybrid proteogenomic metric, QPI-PSI. We then evaluated the ability of these metrics to predict TTN-TV penetrance (in a case-control cohort of 2,446 cases and 922 controls) and AHF expressivity across two independent TTN cardiomyopathy cohorts (Cedars-Sinai cohort, n=128; DCM Precision Medicine cohort, n=180).

**Results:** Multiple TTN transcript isoforms are expressed in human myocardium. PSI-DCM-LR-15 values differed substantially from original PSI calculations (PSI-Original) within the Z-disc and I-band. Proteomic profiling revealed lower TTN peptide abundance in controls compared to DCM. The hybrid metric, QPI-PSI, significantly improved discrimination of TTN-TV penetrance compared to PSI-Original (AUROC 0.694 vs. 0.674, p=0.002). For AHF expressivity, QPI-PSI outperformed PSI-Original in both the Cedars-Sinai cohort (overall AUROC 0.590 vs. 0.562, p=0.304) and the DCM Precision Medicine cohort (overall AUROC 0.587 vs. 0.516, p=0.018). In combined multivariable modeling (n=308), QPI-PSI achieved significantly higher discrimination for AHF than PSI-Original (AUROC 0.639 vs. 0.600, p=0.019), particularly in women (AUROC 0.675 vs. 0.412, p=0.028).

**Conclusions:** The novel hybrid proteogenomic metric QPI-PSI more accurately predicts cardiomyopathy penetrance in TTN-TV carriers and AHF expressivity in patients with TTN cardiomyopathy, particularly among women. With continued validation, these values can help clinical risk stratification in TTN-TV carriers and patients with TTN cardiomyopathy.

## Introduction

Truncating variants in the titin (TTN-TV) gene, comprising stop gain, frameshift, or splice donor/acceptor mutations, are the most common genetic cause of dilated cardiomyopathy (DCM), being identified in 10%-20% of patients ^1^. Cardiomyopathy due to *TTN* truncating variants (TTN-CM) is among the most responsive to heart failure medical therapy, with left ventricular (LV) reverse remodeling observed in 50-60% of cases ^2–4^. However, TTN-CM is also the most frequently identified genetic cause of end-stage heart failure requiring heart transplantation (HT) ^5^.

The metatranscript of the *TTN* gene has 363 coding exons which in turn encode the 35,991 amino acid protein that connects the M-band to Z-disc of the sarcomere ^6^. In addition to its well-established contribution to the passive tension of the cardiomyocyte ^7^, *TTN* directly tunes active contraction by driving length-dependent activation of thick and thin filaments ^8^, functions as a molecular force sensor ^9^, and regulates cardiac protein turnover, hypertrophic growth, and gene expression ^10^. *TTN* is transcribed into 15 isoforms (as per Gencode), with the predominantly expressed isoforms in adult cardiomyocytes being N2BA and N2B ^7,11^. However, the shorter isoforms, Novex-3 ^12^ and Cronos ^13^, also have important functions in both fetal and adult cardiomyocytes. The function of the other 11 transcriptional isoforms remains unclear.

TTN-TVs can be identified in up to 1-3% of the population ^11,14,15^. Prior studies have shown that patients with DCM carry a preponderance of TTN-TVs located in the A-band ^11,16^. The mRNA expression levels of transcribed *TTN* exons within 3 main isoforms (*TTN* N2BA, N2B, and Novex-3) have been used to calculate the “percent spliced in” (PSI), which quantifies the fraction of mRNA transcripts that include a particular exon. Patients with TTN-TVs affecting exons with a PSI > 0.9 (included in >90% of transcripts) are more likely to develop DCM ^11^. As a result, the American College of Medical Genetics and Genomics (ACMG) criteria for variant interpretation in DCM originally required pathogenic TTN-TV to originate in the A-band ^17^. These criteria for pathogenicity were revised to include variants affecting exons outside the A-band with high PSI (PSI > 0.9) ^1^ because there is an association between DCM and TTN-TVs in high PSI exons within the Z-disc, I-band, and M-band. Interestingly, DCM is also associated with TTN-TV in a limited number of low PSI (0.15 to 0.48) exons within the I-band ^18^. Furthermore, there are numerous A-band exons with PSI > 0.9 that have not exhibited significant enrichment in DCM ^18^. These findings suggest that A-band restrictive views of TTN-related DCM may be both scientifically and clinically limiting.

The development of DCM in patients carrying TTN-TV, *i. e.* penetrance, differs from the severity of cardiomyopathy that develops, *i. e.* expressivity ^19,20^. The ability to predict which patients with TTN-CM will have high expressivity and develop advanced heart failure (AHF) requiring specialized therapies including durable left ventricular assist device (LVAD) implantation or HT represents a critical but unmet clinical need. We hypothesized that PSI would predict the risk of AHF in patients with TTN-CM. However, given that PSI values were derived using short-read RNA sequencing and including only 3 of the 15 isoforms, we further hypothesized that recalculating PSI using long-read RNA sequencing and including all 15 *TTN* isoforms would provide more accurate predictions of penetrance and expressivity. Additionally, transcript and protein abundance can be discordant in myocardium ^21^, so we were motivated to develop a method to compare PSI values to exon-specific TTN protein abundance using quantitative peptide mass spectrometry, a value we termed “quantile peptide intensity” (QPI). Finally, we proposed that a hybrid metric, QPI-PSI, combining proteomic and transcriptional exon abundance would improve prediction of the development of TTN cardiomyopathy (penetrance) and its severity (expressivity).

## Methods

### Cardiac Tissue for RNA, Spatial Transcriptomics, and Proteomic Studies

This study was performed in accordance with the ethical principles outlined in the Declaration of Helsinki under protocols approved by the Cedars-Sinai Institutional Review Board (Pro00010979, STUDY00001617, STUDY00004096) and OneLegacy. Informed written consent was obtained from all tissue donors prior to their inclusion in the study. We utilized a total of 66 cardiac tissue samples (Supplemental Table 1) for analysis by 3 methods. For long-read RNA sequencing, samples were obtained from 9 organ donors not utilized for HT (controls) and 25 patients undergoing HT for end-stage DCM. For spatial transcriptomics, samples were obtained from 1 organ donor not utilized for HT and 3 patients undergoing HT for end-stage DCM. For proteomics, samples were obtained from 14 organ donors not utilized for HT (controls)^21^ and 14 patients undergoing HT for end-stage DCM.

### Genetic Testing of Tissue Donors and Cedars-Sinai TTN-CM Cohort

Genetic testing was performed using commercial vendors or via whole exome sequencing as part of a biobank study as previously described ^22^. All genes classified by ClinGen as having a definitive, strong, or moderate association for DCM were tested (Supplemental Table 3) ^23^. Variant consequence and genomic position were determined using Ensembl Variant Effect Predictor ^24^ based on the *TTN* metatranscript (ENST00000589042.5). Variant interpretation was based on that of ClinVar (if the variant was present in ClinVar with an aggregate review status of 2 or more stars) and Franklin by Qiagen if not present in ClinVar ^25^. Genetic testing results for cardiac tissue donors are provided in Supplemental Tables 2.

### Cedars-Sinai Titin Cardiomyopathy Cohort

We identified 132 patients from 124 families that presented to the Advanced Heart Disease or Cardiogenetics clinics at Cedars-Sinai Medical Center between 2012 and 2026 for TTN-CM. In addition, we identified patients through search of the Cedars-Sinai electronic medical record via Deep6 AI ^26^. We defined TTN-CM as: 1) left ventricular ejection fraction (LVEF) < 50% by echocardiography, with or without LV dilation (as defined below), and 2) the presence of a likely pathogenic (LP) variant, pathogenic (P) variant, or variant of undetermined significance in the *TTN* gene. Genetic testing results for the Cedars-Sinai TTN-CM cohort are provided in Supplemental Table 4. AHF was defined as HT or death due to HF. We excluded implantation of an LVAD as AHF because: (1) criteria for LVAD implantation are variable^27^ and may be based on patient characteristics such as body size and blood type alone^28^, and (2) patients with TTN-CM may exhibit significant reverse remodeling after LVAD^29^. In patients who did not have AHF and who presented with an initial LVEF < 40%, LV reverse remodeling was defined as improvement of EF from first to last available echocardiogram of ≥ 10% or achievement of an ejection fraction of ≥ 40% ^30^. We excluded 1 patient with a deep intronic variant and 3 patients with other causes for CM (1 amyloid, 1 ischemic, and 1 with LP variant in another DCM gene).

### Dilated Cardiomyopathy Precision Medicine Cohort

The design ^31^, patient characteristics and genetics ^1^ of the Dilated Cardiomyopathy Precision Medicine (DCM-PM) study have been previously reported. The DCM-PM study was performed in accordance with the ethical principles outlined in the Declaration of Helsinki under protocols approved by the Institutional Review Boards at the Ohio State University and at all clinical sites. All participants gave written informed consent. Due to differences in variant interpretation between the Cedars-Sinai TTN-CM cohort and DCM-PM cohort (specifically the requirement for TV-containing exons to have a PSI > 0.9 to meet criteria for LP/P in DCM-PM) we defined TTN-CM for this cohort as: 1) left ventricular ejection fraction (LVEF) < 50% by echocardiography, 2) left ventricular dilation as ≥ 95^th^ percentile for sex/height, and 3) the presence of a variant of undetermined significance (VUS), LP, or P variant in the *TTN* gene. AHF was again defined as the need for LVAD implantation or HT. We excluded 2 patients due to the presence of an additional cause of cardiomyopathy: 1 with a LP variant in *FLNC* and 1 with a LP variant in *TCAP*.

### Long-read RNA Sequencing, Bioinformatics, and Quality Assessment

RNA was isolated from explanted cardiac tissue using Zymo RNA extraction kit (Zymo Research). RNA purification was performed using columns (Qiagen). Library preparation was performed using the Kinnex RNA kit (PacBio). Parameters of RNA libraries are detailed in Supplementary Table 5. Sequencing was performed using a SMRT cell on a Revio system. The resulting BAM files were mapped to the transcriptome (Gencode version 48) using pbmm2, a PacBio-specific implementation of minimap2 ^32^. The transcriptome was modified to include the *TTN* isoform Cronos which originates at a transcriptional start site in the intron between exon 239 and 240 of the *TTN* metatranscript ^13^. We assessed for novel *TTN* isoforms using “Bambu” ^33^. Detailed methods for the assessment of long-read sequencing quality for *TTN* of are described in the Supplemental Material. Transcripts were quantified using Oarfish ^34,35^. Counts were compared between samples using DESeq2 ^36^.

### Short-read RNA Sequencing and Quality Assessment

Samples were obtained from 56 patients undergoing HT for end-stage DCM. RNA was isolated and purified as above. Library preparation was performed using xGen Broad-Range RNA Library Prep kit (IDT). Parameters of RNA libraries are detailed in Supplementary Table 9. RNA sequencing was performed using NovaSeq 6000 (Illumina). Transcripts were quantified using STAR ^37^. EJ coverage was assessed and compared to long-read coverage using a bespoke R script and results are provided in Supplementary Table 10.

### Spatial Transcriptomics

Detailed methods are described in the Supplemental Material. Briefly, formalin-fixed paraffin-embedded slides from 4 heart explants were obtained: 1 from an organ donor not utilized for HT, 1 from a patient undergoing HT for end-stage DCM due to a *DSP* variant, and 2 patients undergoing HT for end-stage DCM due to a TTN-TV. Slides were prepared and analyzed using the Xenium system (10x Genomics). Probe sequences for TTN are provided in Supplemental Table 11.

### Calculation of Percent Spliced In

We tabulated EJ for all *TTN* isoforms in Gencode version 48 with the addition of Cronos (Supplemental Table 12). Junction sequences 20 base pairs in length (10 bp from each exon) were counted in a sample-wise fashion using Bioconductor ^38^ Biostrings ^39^. PSI was calculated for the *TTN* metatranscript per the original definition ^40^:

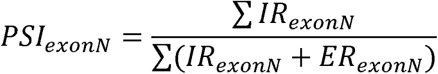

where N=exon number, IR=inclusion reads, ER=exclusion reads. The median PSI value for each exon among all samples was utilized.

### Mass Spectrometry-based Proteomics

Detailed methods are described in the Supplemental Material. Briefly, explants from 28 cardiac tissue samples (n=14 control, n=14 DCM) were fractionated into cytosolic, myofilament, and insoluble protein fractions, trypsin-digested, and analyzed in triplicate via LC-MS/MS in data-independent acquisition (DIA) mode. Peptide identification and quantification were performed using DIA-NN ^41^ against a spectral library constructed from the whole proteome and the 15 human titin *TTN* isoforms. To evaluate isoform-specific expression, *in silico* tryptic digests of the 15 *TTN* isoforms were filtered for unique peptides and mapped against the experimentally detected peptides.

### Calculation of Quantile Peptide Intensity

The protein sequence of the canonical *TTN* metatranscript was digested *in silico* with trypsin, allowing up to one missed cleavage. To eliminate ambiguous mapping, analysis was restricted to exon-unique and junction-unique peptides (peptides mapping to a single exon or a single exon junction [EJ]). All experimentally identified peptides with an intensity greater than 0 in at least one sample were matched to their corresponding exon(s). For peptides spanning an EJ, signal intensities were fractionally apportioned between the 2 constituent exons in proportion to the number of amino acids encoded by each exon. Peptide intensities were summed for each exon and log_2_ transformed. The median exon-specific peptide intensity value among control (n=14) samples was then ranked as a quantile within the context of the full protein. We termed this exon-specific metric “quantile peptide intensity” (QPI).

### Integration of Proteomic and Transcriptomic Metrics (Hybrid QPI-PSI Metric)

To maximize exon-level quantification coverage across the *TTN* gene, we developed a hybrid metric that integrates proteomic-level Quantile Peptide Intensity (QPI) with transcript-level Percent Spliced In (PSI). For exons located outside the A-band, QPI was prioritized whenever peptide signal was detected (QPI > 0). In cases where proteomic data were absent (QPI = 0), we substituted the transcript-level PSI. Additionally, for all exons within the A-band region, PSI was exclusively utilized to account for the high level of expression of the exons in this domain.

### Statistical Analysis

All analysis was performed using R ^42^. All figures were made using “ggplot2” ^43^. PSI values were compared using symmetric mean absolute percentage error (sMAPE) ^44^; correlation coefficient cannot be used since all PSI values of the A-band are equal to 1. sMAPE is given by the formula:

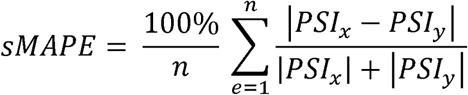

Where PSI_x_ and PSI_y_ represent the values of two different PSI metrics for each exon and n represents the total number of exons. sMAPE values range between 0 and 100%. Univariate logistic modeling with AHF as the response variable was performed using a binomial generalized linear model with a logit link. Given that women were underrepresented in the study population (28% women in the Cedars-Sinai TTN-CM Cohort [Table 3] and 36% women in the DCM Precision Medicine Cohort [Table 6]), inverse probability sample-weighting (w=3 for women and w=1 for men) was implemented for univariate modeling. This weighting scheme accounts for sex-based disparity in TTN-CM penetrance and expressivity and aligns with NIH guidance on Sex as a Biological Variable to ensure equal contribution of male and female profiles to model outcomes ^45^. Variables significant (p < 0.05) in univariate logistic models were included in multivariable models. Multivariable logistic modeling with AHF as the response variable was performed using “glmnet” ^46^ with 10-fold cross-validation and α=0.5 (elastic-net). Inverse probability sample-weighting for women was not implemented for multivariable models. Coefficients were determined at the minimum λ. Prediction performance was assessed using leave-one-out cross validation.

The performance of PSI metrics as single variables and as part of a multivariable model were assessed using the following techniques: (1) model discrimination ^47^ was assessed using the area under the receiver operating characteristic curve (AUROC) and AUROC were compared via the DeLong test in “pROC” ^48^, (2) model calibration^49^ was assessed using the Hosmer–Lemeshow goodness-of-fit test in “ResourceSelection” and plotted using a calibration plot, and (3) model reclassification was assessed using continuous net reclassification index^50^ in ”PredictABEL”^51^.

Detailed methods for the development of the TTN Annotator application are described in the Supplemental Material.

## Results

### TTN Transcript Isoform Quantification

We quantified *TTN* mRNA transcript isoforms using heart tissue collected from 34 donors: 9 organ donors not utilized for HT, 13 patients with end-stage DCM undergoing HT without a TTN-TV (non-TTN-CM), and 12 patients with end-stage DCM undergoing HT due to a TTN-TV (TTN-CM). Among the 13 patients with non-TTN-CM, 6 had disease-causing variants in other genes associated with DCM, specifically *DSP*, *FLNC*, *LMNA*, and *PLN* (Supplemental Table 2). In the 12 patients with TTN-DCM, all *TTN* variants were located in exons with PSI of 1.0 (Supplemental Table 2).

To assess *TTN* transcript isoform expression levels, we quantified long-read sequencing-derived RNA expression levels of all 15 *TTN* transcript isoforms curated in Gencode, as well as Cronos, which was manually curated, with long-read RNA sequencing. We did not identify any novel *TTN* transcript isoforms. The top 6 most highly expressed isoforms were N2BA, N2B, TTN-208, Novex-3, TTN-215, and N2A (Table 1). The *TTN* metatranscript, which is the default *TTN* transcript as defined by the Matched Annotation from NCBI and EMBL-EBI (MANE) project, was in the lower half of *TTN* transcript isoforms by expression levels. Given that this transcript is used for variant interpretation, it is notable that it is transcribed at low levels. Cronos was the transcript with the lowest expression.

**Table 1:** Comparison of *TTN* transcript isoform expression between unused donor hearts (Control) and explants with dilated cardiomyopathy (DCM)

| Transcript | Expression Level |  | Log2 Fold Change | P-value | FDR |
|---|---|---|---|---|---|
|  | Control<br>n=9 | DCM<br>N=25 |  |  |  |
| TTN-213/N2BA | 92510 | 68428 | -0.654 | 0.013 | 0.070 |
| TTN-210/N2B | 13843 | 21270 | 0.807 | 0.020 | 0.090 |
| TTN-208 | 9609 | 4868 | -1.055 | 0.007 | 0.043 |
| TTN-204/Novex-3 | 5393 | 3852 | -0.524 | 0.018 | 0.085 |
| TTN-215 | 4788 | 3105 | -0.955 | 0.000 | 0.006 |
| TTN-202/N2A | 3708 | 1042 | -1.185 | 0.016 | 0.077 |
| TTN-206 | 2333 | 1430 | -0.227 | 0.756 | 0.876 |
| TTN-207 | 937 | 313 | -0.298 | 0.592 | 0.769 |
| TTN-212/Meta | 640 | 783 | 0.750 | 0.530 | 0.722 |
| TTN-205 | 465 | 473 | -0.449 | 0.206 | 0.411 |
| TTN-209 | 259 | 88 | 1.137 | 0.278 | 0.494 |
| TTN-214 | 138 | 153 | 0.270 | 0.396 | 0.611 |
| TTN-201/Novex-2 | 133 | 92 | -0.375 | 0.365 | 0.582 |
| Cronos | 13 | 20 | 1.184 | 0.054 | 0.176 |

Abbreviations: DCM: dilated cardiomyopathy; FDR: false discovery rate; TTN: titin.
Notes: 1: Expression levels are provided in median normalized counts. 2: P-values were calculated using the negative binomial Wald test. 3: FDR is calculated using the Benjamini-Hochberg (BH) procedure.

We compared *TTN* mRNA transcript isoform expression levels between unused donor hearts (controls) and heart explants with DCM, including both non-TTN-CM and TTN-CM (Table 1). Two *TTN* isoforms differed significantly between the groups (false discovery rate [FDR] < 0.05): TTN-208 and TTN-215 were both expressed at lower levels in DCM. Additionally, 4 *TTN* isoforms showed borderline significance (FDR 0.05–0.10), with N2BA, Novex-3, and N2A expressed at lower levels in DCM, and N2B expressed at higher levels. We also compared *TTN* mRNA transcript isoform expression levels between non-TTN-CM and TTN-CM heart explants and found no significant differences in isoform expression (Supplemental Table 8). Taken together, these results show: (1) multiple *TTN* transcript isoforms are expressed in humans, (2) mRNA isoform expression is mostly similar in DCM as compared to normal tissue with the exception of 2 isoforms, and (3) mRNA isoform expression is similar in non-TTN-CM as compared to TTN-CM.

### Spatial Transcriptomics

We used spatial transcriptomics to validate the *TTN* isoform expression levels we measured using long-read RNA sequencing. We performed spatial transcriptomics using the Xenium platform on heart tissue collected from 4 donors: 1 organ donor, 1 patient that underwent HT for DCM due to a *DSP* disease-causing variant, and 2 patients that underwent HT for DCM due to TTN-CM. We developed isoform-specific probes for highly expressed *TTN* isoforms that contain unique splice junctions: N2BA, Novex-3, and metatranscript. For all 4 donors, we found that the count ratio of *TTN* Novex-3 to N2BA and meta-transcript to N2BA were similar between those derived from long-read RNA sequencing and spatial transcriptomics (Figure 1 and Supplemental Table 9). These results validate the accuracy of our *TTN* isoform quantification obtained by long-read RNA sequencing.

**Figure 1:**
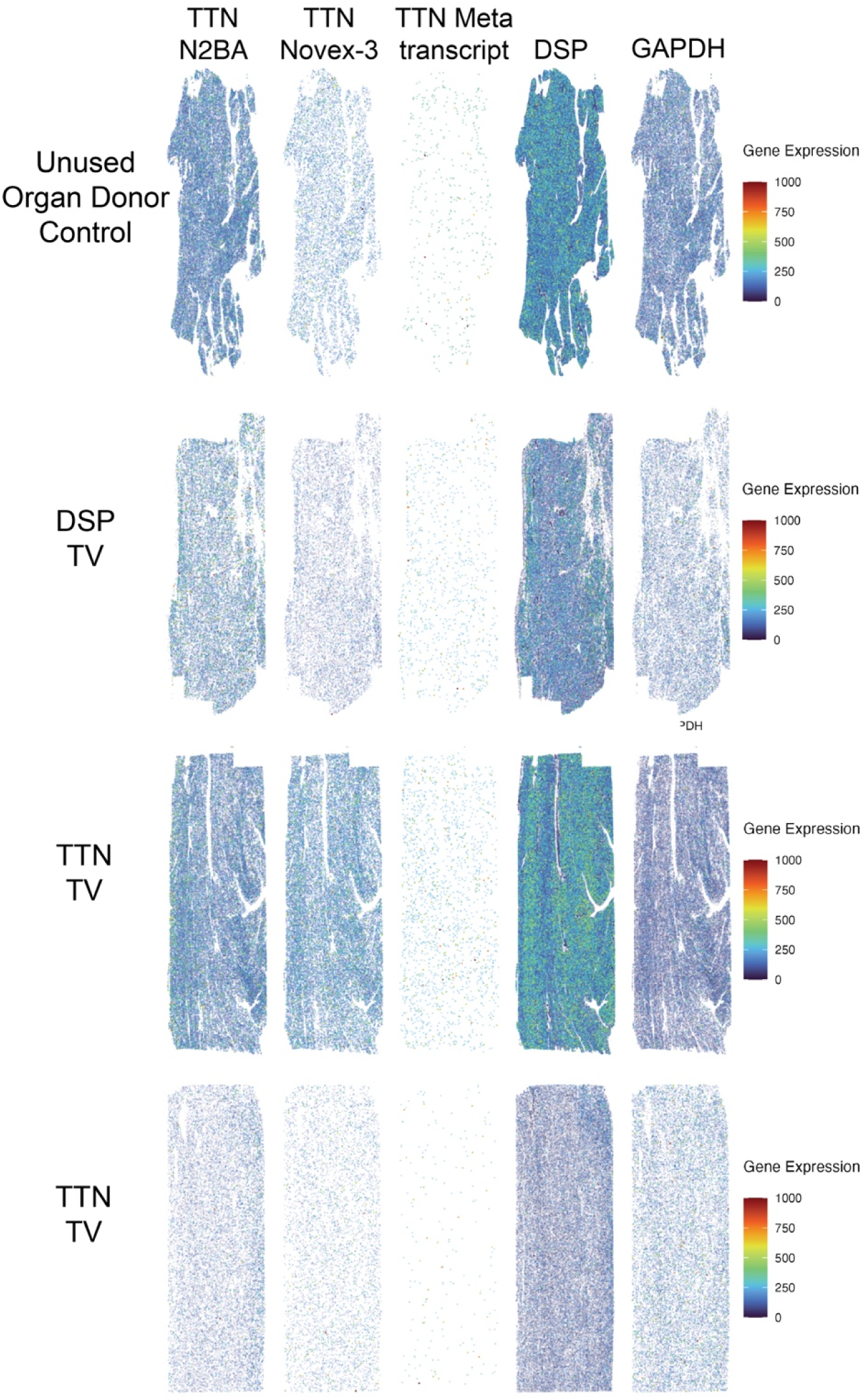
Spatial transcriptomics using explanted heart tissue obtained from an unused organ donor, patient with DCM due to a *DSP* truncating variant, and two patients with DCM due to *TTN* truncating variants. Using probes specific for the N2BA, Novex-3, and metatranscript *TTN* isoforms, we found that the ratio of Novex-3:N2BA and metatranscript:N2BA was similar between all four donors and yielded similar values to those measured using long-read RNA sequencing.

### Calculation of PSI Metrics

We quantified EJ expression for all junctions present in *TTN* transcript isoforms in Gencode. First, we compared EJ expression for the *TTN* metatranscript between the 9 controls and 25 heart explants with DCM and found that the counts of only 4 EJ differed significantly between the groups (false discovery rate [FDR] < 0.05): exons 151-152, 152-153, 156-157, and 219-220 exhibited higher expression in controls. These 4 junctions are in the I-band. Next, calculated exon-specific PSI (based on exons present in the *TTN* metatranscript) for each sample, and found the median value across samples. We hypothesized that a PSI metric calculated using long-read RNA sequencing and including all 15 *TTN* isoforms, which we termed PSI-DCM-LR-15, would provide more accurate predictions of penetrance and expressivity. As expected based on observations of PSI-Original, median PSI-DCM-LR-15 values varied by domain of *TTN* (Figure 2A), with values of 1 for all exons in the A- and M-bands, and values of <1 for 11/28 (39%) and 176/223 (79%) for exons in the Z-disc and I-band.

**Figure 2:**
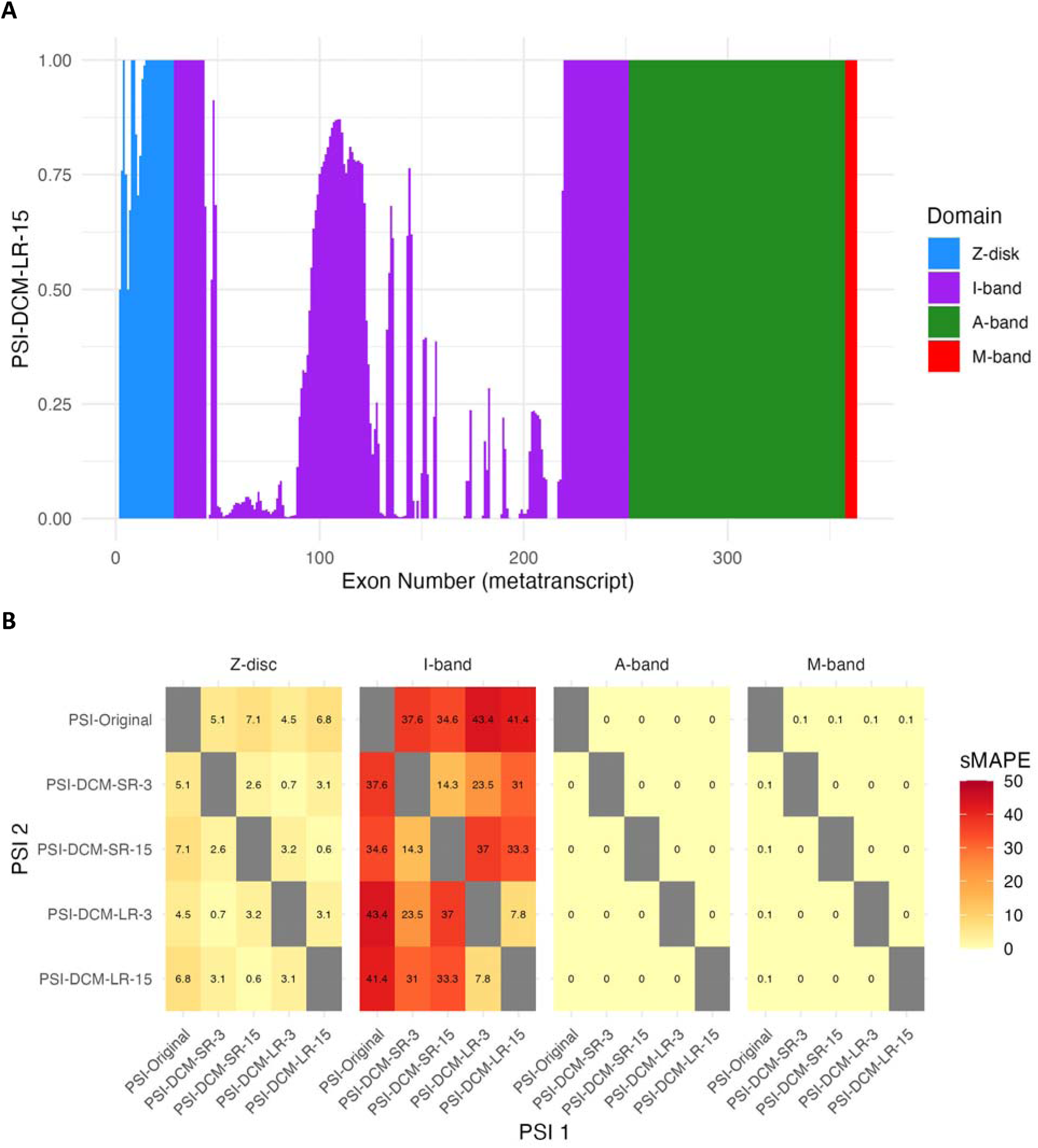
(A) Plot of PSI-DCM-LR-15 values against exon number in the *TTN* metatranscript and colored by *TTN* domain. (B) Heatmap of symmetric mean absolute percent error (sMAPE) values between percent spliced in (PSI) metrics for each domain of the TTN protein. sMAPE values range between 0% and 100%. sMAPE values were highest for the I-band and Z-disc and near 0Q% for the A-band and M-band.

We calculated 3 additional versions of PSI to determine potential sources of variation: 1) PSI-DCM-SR-15, calculated from DCM samples, quantified using short-read RNA sequencing, considering 15 *TTN* isoforms, 2) PSI-DCM-SR-3, calculated from DCM samples, quantified using short-read RNA sequencing, considering 3 *TTN* isoforms (N2BA, N2B, Novex-3), and 3) PSI-DCM-LR-3, calculated from DCM samples, quantified using long-read RNA sequencing, considering 3 *TTN* isoforms. We also compared these PSI values to those calculated by Roberts et al. (PSI-Original) ^11^.

We compared these 5 PSI metrics for the 4 domains of the TTN protein: Z-disc, I-band, A-band, and M-band (Figure 2B and Supplemental Table 15). For the Z-disc, the 5 PSI metrics varied by 0.6% to 7.1%. PSI-DCM-LR-15 differed from PSI-DCM-SR-15 by 0.6% (sMAPE 0.6%), from PSI-DCM-LR-3 by 3.1%, and from PSI-Original by 6.8%. In contrast, PSI-Original differed from PSI-DCM-SR-3 by 5.1%. For the I-band, the 5 PSI metrics varied by 7.8% to 43.4%. PSI-DCM-LR-15 differed from PSI-DCM-SR-15 by 33.3%, from PSI-DCM-LR-3 by 7.8%, and from PSI-Original by 41.4%. In contrast, PSI-Original differed from PSI-DCM-SR-3 by 37.6%. The 5 PSI metrics were identical for the A-band (sMAPE 0%). Lastly, for the M-band, the 5 PSI metrics were nearly identical (sMAPE 0% to 0.1%). These results suggest that the difference in values between PSI-DCM-LR-15 and PSI-Original is due to a combination of both the use of long-vs. short-read RNA sequencing and the use of 15 vs. 3 *TTN* isoforms, as well as intrinsically different values for the exons of the I-band.

### Mass Spectrometry-based TTN Peptide Mapping

To analyze *TTN* isoform proteoform expression, we performed mass spectrometry-based proteomics on 28 samples of explanted cardiac tissue: 14 from heart explants with DCM and 14 controls. We obtained excellent proteome coverage for TTN and observed 1,202 unique TTN proteotypic peptides, represented by 1,358 total precursors comprised of redundant TTN peptides with various charge states and/or post-translational modifications. This represents 20% of the total possible proteotypic peptides of TTN generated from all proteoforms (total possible = 6,053).

First, we sought to identify which TTN proteoforms were represented in the 1,202 unique proteotypic TTN peptides. Of the 16 *TTN* transcript isoforms, 15 yield unique amino acid sequences (Supplemental Table 12), as TTN-211 does not contain an open reading frame. The majority (99%, 1,186/1,202 peptides) of the peptides mapped to the *TTN* metatranscript (Figure 3). There were 16 unique proteotypic peptides that mapped exclusively outside the *TTN* metatranscript, which represents 5% of the total possible proteotypic peptides of TTN generated from proteoforms other than the metatranscript (total possible = 327). Of these 16, 14 mapped to Novex-3 (Figure 3).

**Figure 3:**
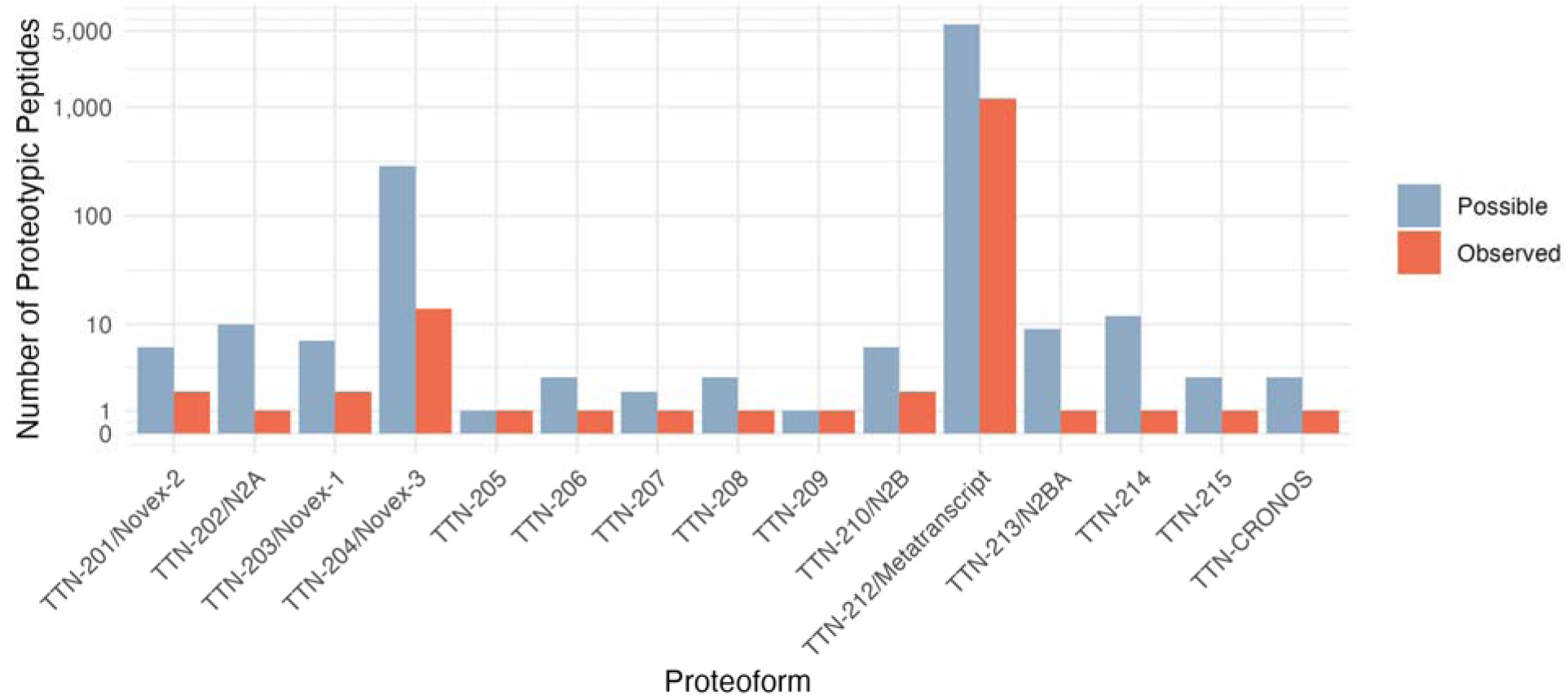
Bar plot of the number of possible (blue) or experimentally-observed (red) peptides from the 15 *TTN* proteoforms. Most experimentally-observed peptides mapped to the TTN-212/*TTN* metatranscript or TTN-204/Novex-3.

Next, we assessed the abundance of peptides translated from *TTN* metatranscript exons 2 to 363. The number of possible proteotypic peptides per exon varied by *TTN* domain, with a median of 9.8, 6.3, 16.5, and 11.5 for the Z-disc, I-band, A-band, and M-band, respectively (Figure 4A). Based on 1,186 peptides that mapped to the *TTN* metatranscript protein product, there were 2/27 exons in the Z-disc (7%), 128/223 exons in the I-band (57%), 3/106 exons in the A-band (3%), and 0/5 exons in the M-band for which we did not identify any peptides (Figure 4B).

**Figure 4:**
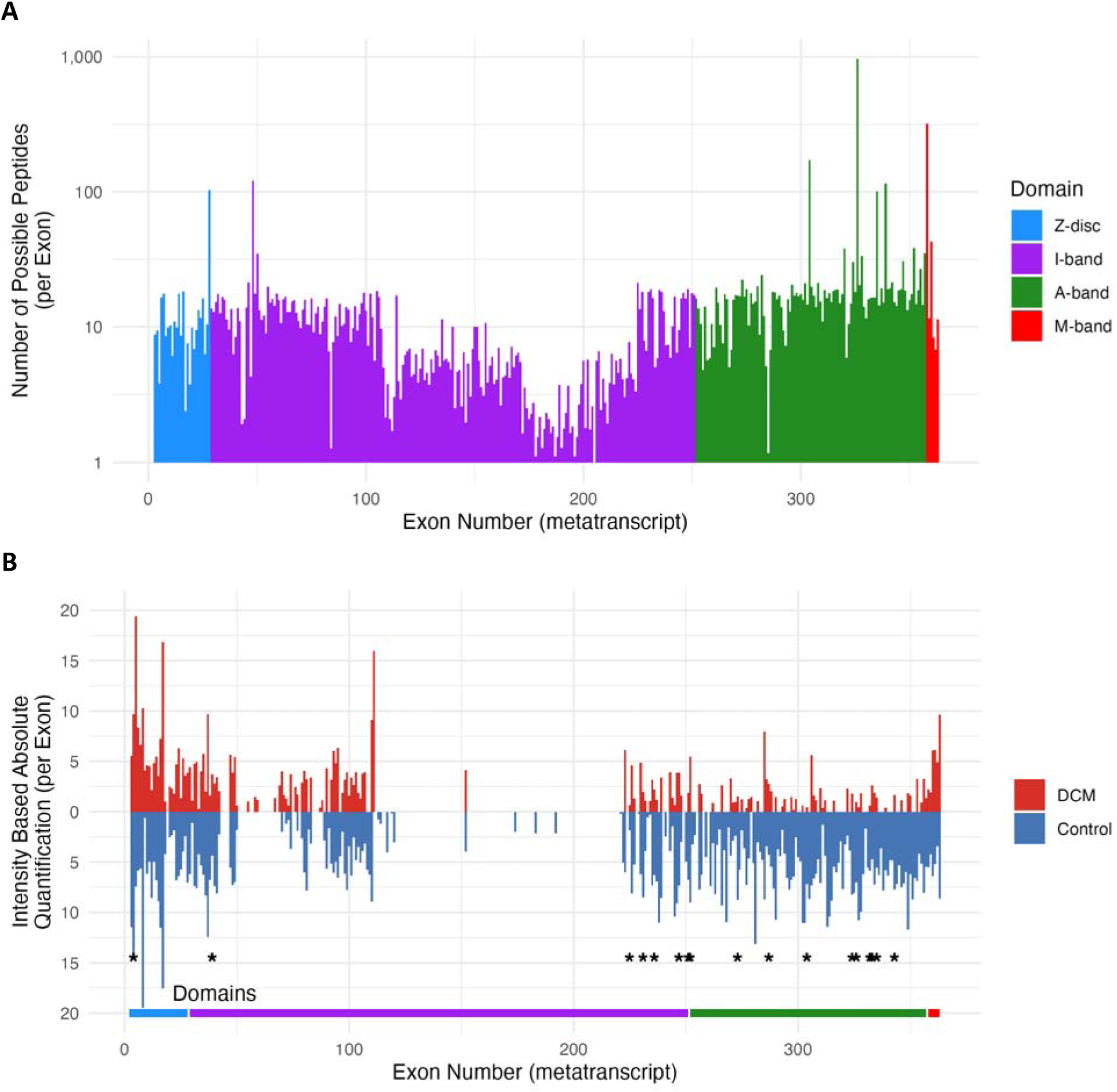
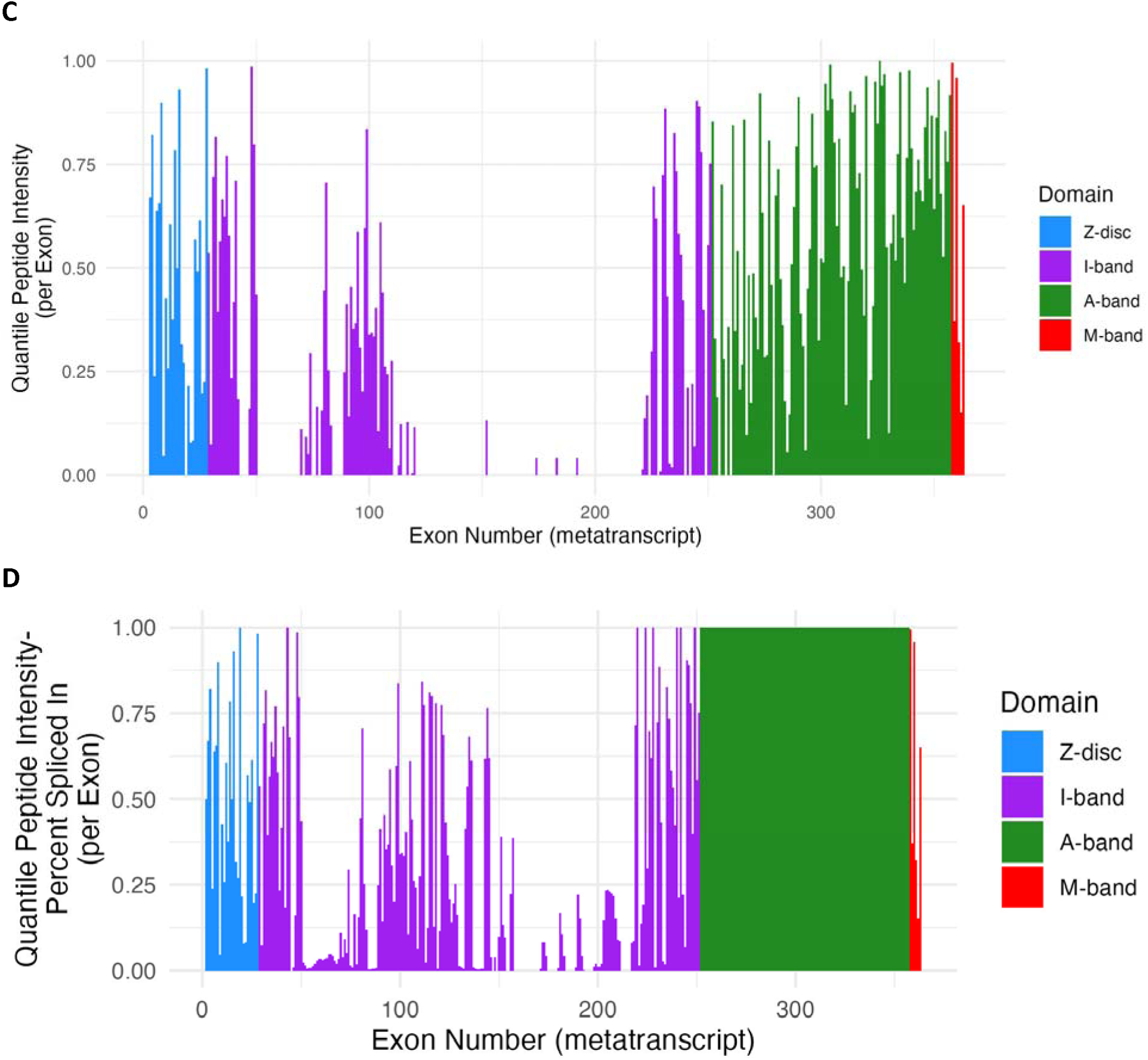
(A) Plot of the possible number of proteotypic peptides for each exon against exon number in the *TTN* metatranscript and colored by *TTN* domain. (B) Plot of Intensity Based Absolute Quantification for each exon against exon number in the *TTN* metatranscript from controls (blue) and DCM explants (red). Asterisks indicate exons with significantly higher expression in controls than DCM. *TTN* domains are colored similar to panel B on the bottom of the plot. (C) Plot of Quantitative Peptide Intensity (QPI) for each exon against exon number in the *TTN* metatranscript and colored by *TTN* domain. (D) Plot of the hybrid metric QPI-percent spliced in (QPI-PSI) for each exon against exon number in the *TTN* metatranscript and colored by *TTN* domain.

We quantified peptide abundance using intensity-based absolute quantification (IBAQ) which accounts for the varying number of possible proteotypic peptides per exon. We compared the exon-specific IBAQ values between the 14 heart explants with DCM and 14 controls (Figure 4B). Mean IBAQ differed by domain and group: 5.6, 1.1, 0.9, and 5.1 for DCM explants in the Z-disc, I-band, A-band, and M-band, respectively, as compared to 6.8, 1.8, 5.8, and 5.6 for controls (p<0.001 by ANOVA). We found that the iBAQ of 1 Z-disc exon (exon 4), 6 I-band exons (exons 39, 225, 231, 236, 247, 251), and 10 A-band exons (exons 252, 273, 287, 304, 324, 326, 332, 333, 335, 343) were significantly higher in controls as compared to DCM. There were no M-band exons that differed significantly. We then ranked each exon’s sum intensity as a quantile (considering all exons in the *TTN* metatranscript). We termed this exon-specific metric “quantile peptide intensity (QPI).” We found that QPI varied substantially between the domains of TTN: the median QPI was 0.49, 0.34, 0.64, and 0.51 for the Z-disc, I-band, A-band, and M-band, respectively (Figure 4C). Because there were 144 exons with missing values for QPI (no peptides detected), we also developed a hybrid metric that we termed QPI-PSI that that integrates proteomic-level QPI with transcript-level PSI (Figure 4D and Supplemental Table 16).

Taken together, these protein-level data suggest several findings: (1) most experimentally-identified TTN peptides map to the protein product of the canonical *TTN* metatranscript, (2) a small number of TTN peptides map to the non-canonical isoform Novex-3, and (3) proteoform/peptide abundance is lower in DCM than in controls.

### Predictors of Penetrance in TTN Cardiomyopathy

We sought to determine the potential of exon abundance as measured using PSI, QPI, and QPI-PSI to predict penetrance of TTN-TV. We utilized data from Vatta et al. ^18^ who identified in 2,446 TTN-TV cardiomyopathy/arrhythmia cases referred for genetic testing and 922 TTN-TV in gnomAD v3.1. We performed case-control analysis using a logistic model. We found that all four PSI metrics, PSI-Original, PSI-DCM-LR-15, QPI, and QPI-PSI were associated with an increased odds of an exon containing cases (Table 2). Discrimination, as measured by the AUROC, was significantly higher for QPI-PSI as compared to PSI-Original (0.694 vs. 0.674; p=0.002) and PSI-DCM-LR-15 (0.694 vs. 0.674; p=0.002).

**Table 2:**
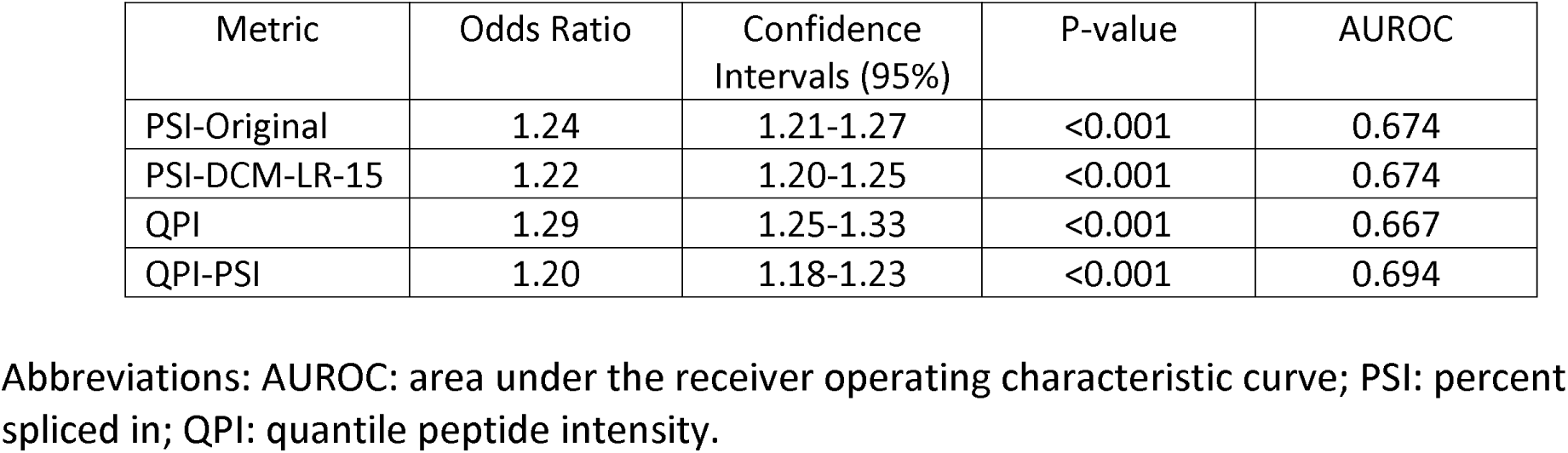
Performance of exon abundance metrics for penetrance of titin truncating variants

| Metric | Odds Ratio | Confidence Intervals (95%) | P-value | AUROC |
|---|---|---|---|---|
| PSI-Original | 1.24 | 1.21-1.27 | <0.001 | 0.674 |
| PSI-DCM-LR-15 | 1.22 | 1.20-1.25 | <0.001 | 0.674 |
| QPI | 1.29 | 1.25-1.33 | <0.001 | 0.667 |
| QPI-PSI | 1.20 | 1.18-1.23 | <0.001 | 0.694 |

Abbreviations: AUROC: area under the receiver operating characteristic curve; PSI: percent spliced in; QPI: quantile peptide intensity.

**Table 3:** Characteristics, echocardiographic and genetic parameters of patients in the Cedars-Sinai TTN Cardiomyopathy Cohort based on whether they developed advanced heart failure

| Characteristic | TTN-CM Without AHF<br>n=88 | TTN-CM With AHF<br>n=40 | P-value |
|---|---|---|---|
| Age of CM onset | 49.0 [36.7;59.8] | 46.0 [37.1;51.9] | 0.190 |
| Sex: female | 29 (33.0%) | 7 (17.5%) | 0.112 |
| Race |  |  | 0.084 |
| Asian | 9 (10.2%) | 3 (7.50%) |  |
| Black | 8 (9.09%) | 9 (22.5%) |  |
| Other | 5 (5.68%) | 5 (12.5%) |  |
| White | 66 (75.0%) | 23 (57.5%) |  |
| HTN | 31 (35.2%) | 17 (42.5%) | 0.555 |
| DM | 13 (14.8%) | 12 (30.0%) | 0.076 |
| SBP at presentation | 117 [108;127] | 106 [95.5;112] | <0.001 |
| Family history of CM | 46 (52.3%) | 24 (61.5%) | 0.438 |
| Atrial fibrillation | 35 (39.8%) | 21 (52.5%) | 0.249 |
| Ventricular tachycardia | 22 (25.0%) | 28 (70.0%) | <0.001 |
| *Echocardiographic parameters* |  |  |  |
| Echo at presentation |  |  |  |
| EF | 27.2 [17.8;40.2] | 20.0 [15.0;27.5] | 0.008 |
| LVIDD | 58.5 (8.32) | 65.5 (11.4) | 0.004 |
| Echo at last follow up |  |  |  |
| EF | 44.5 [35.0;53.0] | 16.0 [14.5;20.0] | <0.001 |
| LVIDD | 56.0 [50.0;60.0] | 65.0 [60.0;72.2] | <0.001 |
| Change in EF | 9.00 [3.00;23.8] | -2.00 [-8.00;0.50] | <0.001 |
| LVRR | 41 (47.7%) | 1 (2.56%) | <0.001 |
| *Genetic parameters* |  |  |  |
| Location of TTN-TV |  |  | 0.075 |
| Z-disc | 7 (7.95%) | 0 (0.00%) |  |
| I-band | 19 (21.6%) | 5 (12.5%) |  |
| A-band | 62 (70.5%) | 35 (87.5%) |  |
| TTN-TV consequence |  |  | 0.216 |
| frameshift | 36 (40.9%) | 23 (57.5%) |  |
| stop-gain | 40 (45.5%) | 14 (35.0%) |  |
| canonical splice | 11 (12.5%) | 2 (5.00%) |  |
| other | 1 (1.14%) | 1 (2.50%) |  |

Abbreviations: AHF: advanced heart failure; CM: cardiomyopathy; DM: diabetes mellitus; EF: ejection fraction; HTN: hypertension; LVAD: left ventricular assist device; LVIDD: left ventricular internal diastolic diameter; LVRR: left ventricular reverse remodeling; SBP: systolic blood pressure; TTN-TV: titin truncating variant.
Notes: P-values were calculated using Student’s t-test for continuous normal variables, Wilcoxon rank-sum test for continuous non-normal variables, and chi-squared test for categorical variables.

### Clinical Relevance of PSI Metrics in the Cedars-Sinai TTN-CM Cohort

We next determined the clinical relevance of the QPI-PSI metric for 128 patients in the Cedars-Sinai TTN Cardiomyopathy Cohort. All patients had been treated with guideline-directed medical therapy. During the study period 40 (31%) patients developed AHF, as defined by HT or death due to HF. Characteristics of TTN-CM patients without and with AHF are provided in Table 3. Significant differences between the non-AHF and AHF groups included higher systolic blood pressure (117 vs. 106 mmHg, p<0.001), lower frequency of ventricular tachycardia (25% vs. 70%, p<0.001), higher LVEF at presentation (27% vs. 20%, p=0.008) and smaller left ventricular internal diastolic dimension at presentation (LVIDD; 59 vs 66 mm, p=0.004). At their last follow up echocardiogram at a median of 2.8 years later (interquartile range 0.9-6.4 years) the EF was higher (45% vs. 16%, p<0.001) and LVIDD was lower (56 vs. 65 mm, p<0.001) in the non-AHF group. There were significantly more patients in the non-AHF group that experienced left ventricular reverse remodeling (48% vs. 3%, p<0.001).

We performed a univariate logistic model with AHF as the endpoint and 11 potential predictor variables including the 4 PSI metrics using inverse probability weighting to account for the distribution of women (Table 4). We found that increasing age of CM onset, female sex, and a non-A-band location for the TTN-TV were associated with a decreased risk of AHF, while black race and a variant consequence of frameshift were associated with an increased risk of AHF. With regards to the PSI metrics, we found that PSI-Original, PSI-DCM-LR-15, and QPI-PSI were associated with a significantly increased risk of AHF. In an unweighted logistic model (Supplemental Table 17), increasing age of CM onset, female sex and mutation consequence of frameshift lost statistical significance, while diabetes gained statistical significance. With regards to the PSI metrics, only QPI-PSI was associated with an increased risk of AHF at borderline statistical significance (p=0.05).

**Table 4:** Results of univariate logistic model for advanced heart failure in the Cedars-Sinai TTN Cardiomyopathy Cohort

| Characteristic | Odds Ratio | 95% Confidence Intervals | P-value |
|---|---|---|---|
| Age of CM onset | 0.758 | 0.604-0.943 | 0.014 |
| Sex: female | 0.432 | 0.225-0.812 | 0.010 |
| Race |  |  |  |
| White | reference | reference | reference |
| Asian/Other | 2.571 | 1.049-6.205 | 0.036 |
| Black | 1.423 | 0.595-3.230 | 0.41 |
| HTN | 1.117 | 0.582-2.114 | 0.737 |
| DM | 1.910 | 0.838-4.227 | 0.114 |
| Location of TTN-TV |  |  |  |
| A-band | reference | reference | reference |
| I-band/Z-disc | 0.196 | 0.065-0.481 | 0.001 |
| TTN-TV consequence |  |  |  |
| stop-gain | reference | reference | reference |
| frameshift | 2.051 | 1.044-4.162 | 0.041 |
| canonical splice | 0.357 | 0.054-1.401 | 0.193 |
| other | 3.750 | 0.143-98.394 | 0.359 |
| PSI Metrics |  |  |  |
| PSI-Original | 1.457 | 1.107-2.441 | 0.040 |
| PSI-DCM-LR-15 | 1.260 | 1.050-1.690 | 0.043 |
| QPI | 1.185 | 0.995-1.433 | 0.067 |
| QPI-PSI | 1.367 | 1.121-1.852 | 0.011 |

Abbreviations: DM: diabetes mellitus; HTN: hypertension; PSI: percent spliced in; QPI: quantile peptide intensity; TTN-TV: titin truncating variant.
Notes: 1: Odds ratio for “Age of CM onset” is per decade. 2: Logistic model performed with inverse probability sample-weighting (weight for women=3, weight for men=1).

Table 5 lists the discrimination of these 4 PSI metrics for AHF in the Cedars-Sinai TTN Cardiomyopathy Cohort. We found that QPI-PSI had a numerically higher AUROC for prediction of AHF that was not statistically significant as compared to PSI-Original (0.590 vs. 0.562; p=0.304). We also compared performance by sex and found that QPI-PSI provided better discrimination for AHF in women than for men with a statistically significant increase in the AUROC as compared to PSI-Original (0.707 vs. 0.603; p=0.007). QPI-PSI provided borderline significant reclassification as compared to PSI-Original, with a continuous net reclassification index of 0.125 (95% CI -0.0188-0.2688, p=0.088).

**Table 5:** Performance of PSI metrics for prediction of advanced heart failure in the Cedars-Sinai TTN Cardiomyopathy Cohort

| Metric | AUROC<br>M/W<br>n=128 | AUROC<br>Men<br>n=92 | AUROC<br>Women<br>n=36 |
|---|---|---|---|
| PSI-Original | 0.562 | 0.545 | 0.603 |
| PSI-DCM-LR-15 | 0.551 | 0.535 | 0.603 |
| QPI | 0.584 | 0.570 | 0.584 |
| QPI-PSI | 0.590 | 0.550 | 0.707 |

Abbreviations: AUROC: area under the receiver operating characteristic curve; AUPRC: area under precision-recall curve; PSI: percent spliced in; QPI: quantile peptide intensity.

**Table 6:** Comparison of characteristics, echocardiographic and genetic parameters of patients in the Cedars-Sinai TTN Cardiomyopathy cohort and the Dilated Cardiomyopathy Precision Medicine cohort

| Characteristic | Cedars-Sinai<br>n=128 | DCM Precision Medicine<br>n=180 | P-value |
|---|---|---|---|
| Age of CM onset | 47.5 [36.8;56.5] | 42.3 [33.7;53.0] | 0.008 |
| Sex: female | 36 (28.1%) | 64 (35.6%) | 0.212 |
| Race |  |  | <0.001 |
| Asian | 12 (9.38%) | 0 (0.00%) |  |
| Black | 17 (13.3%) | 45 (25.0%) |  |
| Other | 10 (7.81%) | 0 (0.00%) |  |
| White | 89 (69.5%) | 135 (75.0%) |  |
| HTN | 48 (37.5%) | 74 (41.1%) | 0.603 |
| DM | 25 (19.5%) | 39 (21.7%) | 0.754 |
| HT | 40 (31.2%) | 40 (22.2%) | 0.099 |
| *Echocardiographic parameters* |  |  |  |
| EF | 21.2 [15.8;35.0] | 20.0 [15.0;27.0] | 0.010 |
| LVIDD | 60.0 [54.0;66.0] | 63.0 [58.0;67.0] | 0.002 |
| *Genetic parameters* |  |  |  |
| Location of TTN-TV |  |  | 0.107 |
| Z-disc | 7 (5.47%) | 6 (3.33%) |  |
| I-band | 24 (18.8%) | 34 (18.9%) |  |
| A-band | 97 (75.8%) | 133 (73.9%) |  |
| M-band | 0 (0.00%) | 7 (3.89%) |  |
| TTN-TV consequence |  |  | 0.010 |
| frameshift | 59 (46.1%) | 73 (40.6%) |  |
| stop-gain | 54 (42.2%) | 61 (33.9%) |  |
| canonical splice | 13 (10.2%) | 29 (16.1%) |  |
| other | 2 (1.56%) | 17 (9.44%) |  |

Abbreviations: AHF: advanced heart failure; CM: cardiomyopathy; DM: diabetes mellitus; EF: ejection fraction; HT: heart transplantation; HTN: hypertension; LVIDD: left ventricular internal diastolic diameter; TTN-TV: titin truncating variant.

**Table 7:** Performance of PSI metrics for prediction of advanced heart failure in a multivariable model

| Metric | AUROC<br>M/W<br>n=308 | AUROC<br>Men<br>n=208 | AUROC<br>Women<br>n=100 |
|---|---|---|---|
| PSI-Original | 0.600 | 0.576 | 0.410 |
| QPI-PSI | 0.639 | 0.617 | 0.675 |

Abbreviations: AUROC: area under the receiver operating characteristic curve; AUPRC: area under precision-recall curve; PSI: percent spliced in; QPI: quantile peptide intensity.

### Clinical Relevance of PSI Metrics in the DCM-PM Study

We then validated the clinical relevance of the QPI-PSI metric in an independent and multicenter cohort of 180 patients with TTN-CM from the DCM-PM Study. We compared the characteristics of the Cedars-Sinai TTN-CM cohort to the DCM-PM cohort (Table 6). The two groups were similar for frequency of AHF (31% vs. 22%, p=0.100), women (28% vs. 36%, p=0.212), hypertension (38% vs. 41%, p=0.603), diabetes mellitus (20% vs. 22%, p=0.754), and in the domain of the TTN protein containing the TTN-TV (p=0.107). Significant differences between the cohorts included an older median age of CM onset in the CS group (48 vs. 42 years, p=0.008), higher LVEF (21% vs. 20%, p=0.010), smaller LVIDD (60 vs. 63 mm, p=0.002), a more diverse racial composition (p<0.001), and a different profile of TTN-TV consequences (p=0.010).

We again performed univariate logistic modeling with AHF as the endpoint and 11 potential predictor variables including the 4 PSI metrics using inverse probability weighting to account for the distribution of women (Supplemental Table 18). We found that women and a non-A-band location for the TTN-TV were associated with a decreased risk of AHF, while hypertension, diabetes mellitus, and variant consequence of frameshift were was associated with an increased risk of AHF. With regards to the PSI metrics, we found that only QPI-PSI was associated with a significantly increased risk of AHF.

Supplemental Table 19 lists the discrimination of these 4 PSI metrics for AHF in the DCM-PM Study cohort. We found that QPI-PSI provided the best discrimination for AHF, and the difference in AUROC was statistically significant as compared to PSI-Original (0.587 vs. 0.516; p=0.018). We also compared performance by sex and found that QPI-PSI provided better discrimination for women than for men with a statistically significant increase in the AUROC as compared to PSI-Original (0.648 vs. 0.546; p<0.001). QPI-PSI provided significant reclassification as compared to PSI-Original, with a continuous net reclassification index of 0.218 (95% CI 0.087-0.349, p=0.001).

### Multivariable Modeling ofAHF Incorporating PSI Metrics

Finally, we performed multivariable modeling to validate the utility of QPI-PSI for prediction of AHF as compared to PSI-Original. Given the small number of events in each cohort, we combined the cohorts for the purposes of multivariable modeling. We trained and tested two a penalized logistic models for AHF using a leave-one-patient-out cross-validation strategy: 1 model using PSI-Original and 1 model using QPI-PSI. In both models, the PSI metric was retained as a variable, *i. e.* was significant in a multivariable model (Supplemental Figure 1). The AUROC of the model containing QPI-PSI (0.639) was significantly higher than that of the model containing PSI-Original (0.639 vs. 0.600; p=0.019; Figure 5A). QPI-PSI provided better discrimination for women than for men with a statistically significant increase in the AUROC as compared to PSI-Original (0.675 vs. 0.412; p=0.028). We also assessed the calibration of the two models and both models provided adequate calibration with p>0.05 by the Hosmer-Lemeshow Goodness of Fit test (PSI-Original p=0.103; QPI-PSI 0.066; Figure 5B).

**Figure 5:**
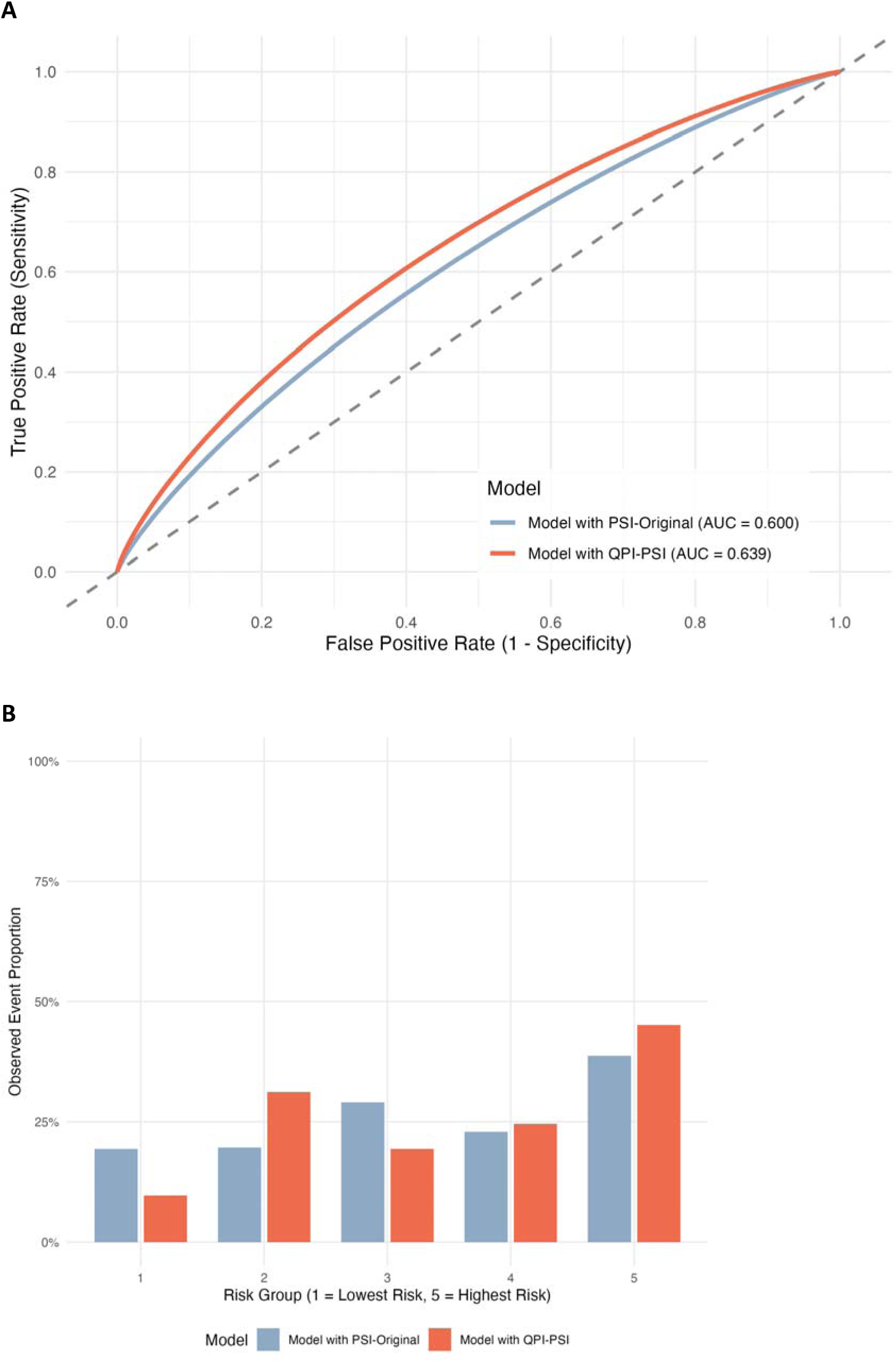
(A) Receiver operator characteristic (ROC) curve for two multivariable logistic models. The area under the ROC curve for the model containing PSI-Original was 0.600 (blue) while the area under the ROC curve for the model containing QPI-PSI was 0.639. The difference between the curves was statistically significant (p=0.019). (B) Calibration curve showing the percentage of the group experiencing advanced heart failure as the predicted probability of risk from the multivariable model containing PSI-Original (blue) or QPI-PSI (red) increases. Both models showed adequate calibration with non-significant p-values by the Hosmer-Lemeshow test.

To facilitate clinical utilization of the QPI-PSI as both a single metric of AHF and as part of a multivariable model, we developed the “TTN Annotator” web application available at https://evanpk-ttn-annotator.share.connect.posit.cloud.

## Discussion

In this study, we first investigated *TTN* mRNA transcript isoform levels using long-read RNA sequencing in explanted cardiac tissue. We found that multiple *TTN* transcript isoforms are expressed in humans, isoform expression is mostly similar in DCM as compared to normal tissue with exception of 2 isoforms, and isoform expression is similar in non-TTN-CM as compared to TTN-CM. We developed additional versions of PSI using long- and short-read RNA sequencing and found that the PSI metrics differed substantially for the Z-disc and I-band domains of TTN, but minimally for the A- and M-bands. We then performed mass spectrometry-based proteomics on explanted cardiac tissue and found that most experimentally-identified TTN peptides map to the protein product of the canonical *TTN* metatranscript, a small number of TTN peptides map to the non-canonial isoform Novex-3, and proteoform/peptide abundance is lower in DCM than in controls. We developed a new quantitative metric for exon abundance at the protein level we termed QPI. Combining QPI and PSI into a hybrid metric, QPI-PSI, improved prediction of penetrance and expressivity (development of AHF) in TTN-CM —especially in women—both as a single variable and within a multivariable model, compared to PSI-Original.

Our results have important clinical implications. When reporting TTNtv, genetic testing laboratories frequently include the PSI value for the exon containing or affected by the variant in the clinical report to provide the ordering clinician with insight into its potential impact. As reported by Roberts *et al.* ^11^, TTNtv affecting exons with high PSI (PSI > 0.9) are more likely to cause TTN-CM and advanced heart failure (AHF), whereas those with low PSI (PSI < 0.9) are more commonly found in healthy individuals. Here, we have shown improved discrimination using QPI-PSI for penetrance and expressivity compared to the original PSI categories developed by Roberts et al. Thus, the exon-specific values of QPI-PSI developed in this study will serve as valuable reference metrics for diagnostic laboratories. Furthermore, while Roberts *et al.* established the foundational relationship between PSI and clinical outcomes, we have advanced this field by demonstrating a continuous relationship between QPI-PSI, penetrance, and expressivity, moving beyond a strictly categorical framework. We have also developed a web application to facilitate use of the QPI-PSI metric and the multivariable prediction model.

We identified several patient characteristics associated with an increased risk of AHF in TTN-CM: male sex, Black race, diabetes mellitus, and frameshift variant consequence. It is well established that men are at increased risk of adverse outcomes in the setting of TTN-CM ^52^. Although TTN-CM is less frequent—and *TTN*-TVs have previously been questioned as a cause of DCM—in individuals of African ancestry ^1,53^, more recent studies have confirmed their pathogenic importance in this population ^54,55^, who also face an overall increased risk of adverse outcomes in DCM ^56^. Likewise, diabetes mellitus increases the penetrance of TTN-TVs ^57^ and is associated with worse outcomes in non-ischemic DCM ^58^. While we found that frameshift TTN-TVs were more strongly associated with AHF, previous research on TTN-TV penetrance found that frameshift carriers were not at an increased risk of developing TTN-CM ^57^; as such, this finding will require replication in larger patient cohorts. Lastly, because our final multivariable model had an AUROC of 0.639 overall, additional research is needed to identify genetic and non-genetic factors associated with the penetrance and expressivity of TTN-TV.

We found that only two *TTN* transcriptional isoforms (*TTN-208* and *TTN-215*) and four exon junctions (EJs) differed between control hearts and DCM explants, all showing higher expression in controls. Furthermore, we observed no differences in *TTN* transcriptional isoforms between non-TTN-CM and TTN-CM, aligning with previous reports ^11,59^. In contrast, proteomic analysis revealed 17 exons that were more abundant in control hearts than in DCM explants, though limited sample sizes prevented a comparison between non-TTN-CM and TTN-CM. Together, these findings suggest that for a highly expressed gene like *TTN*, disease-related changes in DCM may be more pronounced at the protein level rather than the transcriptional level—a phenomenon similarly observed for *MYBPC3* ^60^.

This study has both strengths and limitations. Whereas many prior studies of *TTN* mRNA expression have relied on animal models, a key strength of this work is the examination of explanted heart tissue from human organ donors and patients with TTN-CM, offering direct clinical translation. Nevertheless, only a small number of samples were analyzed, and the QPI and PSI values require replication in larger cohorts. In particular, the penetrance and expressivity of TTN-CM is lower in women, the molecular underpinnings of which remain uncertain. Differences in PSI could contribute to these differences. Indeed, we found that our novel metric of TTN-TV expressivity QPI-PSI improved risk stratification for AHF more in women than in men. Additional work is needed to determine if PSI or QPI-PSI varies by sex. There were a substantial number of *TTN* metatranscript exons that did not yield detectable peptides in our mass spectrometry analysis. This reflects both the sequence repetitiveness of the I-band, which restricts unique proteotypic peptide mapping, and smaller exons that yield less proteotypic peptides. To assess low-abundance exons, future studies utilizing targeted proteomic approaches (such as parallel reaction monitoring) or enrichment strategies will be beneficial. Finally, because the two TTN-CM cohorts were derived from AHF programs and represent a more severely affected patient population, these findings require validation across a broader spectrum of TTN-CM severity.

In conclusion, this study demonstrates that a novel hybrid proteogenomic metric, QPI-PSI, which integrates transcript and protein exon abundance, more accurately predicts cardiomyopathy penetrance in TTN-TV carries and AHF expressivity in patients with TTN-CM. While QPI-PSI remains investigational and necessitates prospective clinical validation, these findings suggest that multi-omics and proteogenomic strategies hold strong potential to improve risk stratification across genetic cardiomyopathies.

## Data Availability

Data will be made available on publication of the manuscript

## Acknowledgements/Funding

This work was funded by seed money support from the Smidt Discovery Fund of the Smidt Heart Institute (E.K.), the American Heart Association (AHA) Postdoctoral Fellowship, AHA Award Number: 829444 (A.B.) and National Institutes of Health (NIH) 1R01HL155346-01A1 (J.V.E.) and 1R01HL144509-01 (J.V.E.). J.V.E. is the Erika Glazer Endowed Chair in Women’s Heart Health. We would also like to acknowledge access to mass spectrometers via the Cedars-Sinai Medical Center Proteomics and Metabolomics Core. Research reported in this publication was also supported by a parent award from the National Heart, Lung, and Blood Institute of the NIH under Aware Number R01HL128857 (R.E.H.), which included a supplement from the National Human Genome Research Institute. The content is solely the responsibility of the authors and does not necessarily represent the official views of the NIH.

## References

1. Jordan E, Kinnamon DD, Haas GJ, Hofmeyer M, Kransdorf E, Ewald GA, et al. Genetic Architecture of Dilated Cardiomyopathy in Individuals of African and European Ancestry. JAMA 2023;330:432–441. doi: 10.1001/jama.2023.11970

2. Jansweijer JA, Nieuwhof K, Russo F, Hoorntje ET, Jongbloed JD, Lekanne Deprez RH, et al. Truncating titin mutations are associated with a mild and treatable form of dilated cardiomyopathy. Eur J Heart Fail 2017;19:512–521. doi: 10.1002/ejhf.673

3. Verdonschot JAJ, Hazebroek MR, Wang P, Sanders-van Wijk S, Merken JJ, Adriaansen YA, et al. Clinical Phenotype and Genotype Associations With Improvement in Left Ventricular Function in Dilated Cardiomyopathy. Circ Heart Fail 2018;11:e005220. doi: 10.1161/CIRCHEARTFAILURE.118.005220

4. Escobar-Lopez L, Ochoa JP, Mirelis JG, Espinosa MA, Navarro M, Gallego-Delgado M, et al. Association of Genetic Variants With Outcomes in Patients With Nonischemic Dilated Cardiomyopathy. J Am Coll Cardiol 2021;78:1682–1699. doi: 10.1016/j.jacc.2021.08.039

5. Kim Y, Gunnarsdottir OB, Viveiros A, Reichart D, Quiat D, Willcox JAL, et al. Genetic Contribution to End-Stage Cardiomyopathy Requiring Heart Transplantation. Circ Genom Precis Med 2023;16:452–461. doi: 10.1161/CIRCGEN.123.004062

6. Furst DO, Osborn M, Nave R, Weber K. The organization of titin filaments in the half-sarcomere revealed by monoclonal antibodies in immunoelectron microscopy: a map of ten nonrepetitive epitopes starting at the Z line extends close to the M line. J Cell Biol 1988;106:1563–1572. doi: 10.1083/jcb.106.5.1563

7. van der Pijl R, Nusayr E, Strom J, Slater R, Gohlke J, Hourani Z, et al. Importance of N2BA Titin in Maintaining Cardiac Homeostasis and Its Role in Dilated Cardiomyopathy. Circ Heart Fail 2025;18:e012083. doi: 10.1161/CIRCHEARTFAILURE.124.012083

8. Ait-Mou Y, Hsu K, Farman GP, Kumar M, Greaser ML, Irving TC, et al. Titin strain contributes to the Frank-Starling law of the heart by structural rearrangements of both thin-and thick-filament proteins. Proc Natl Acad Sci U S A 2016;113:2306–2311. doi: 10.1073/pnas.1516732113

9. Puchner EM, Alexandrovich A, Kho AL, Hensen U, Schafer LV, Brandmeier B, et al. Mechanoenzymatics of titin kinase. Proc Natl Acad Sci U S A 2008;105:13385–13390. doi: 10.1073/pnas.0805034105

10. Lange S, Xiang F, Yakovenko A, Vihola A, Hackman P, Rostkova E, et al. The kinase domain of titin controls muscle gene expression and protein turnover. Science 2005;308:1599–1603. doi: 10.1126/science.1110463

11. Roberts AM, Ware JS, Herman DS, Schafer S, Baksi J, Bick AG, et al. Integrated allelic, transcriptional, and phenomic dissection of the cardiac effects of titin truncations in health and disease. Sci Transl Med 2015;7:270ra276. doi: 10.1126/scitranslmed.3010134

12. Hashimoto K, Ohira M, Kodama A, Kimoto M, Inoue M, Tone S, et al. Loss of connectin novex-3 leads to heart dysfunction associated with impaired cardiomyocyte proliferation and abnormal nuclear mechanics. Sci Rep 2024;14:13727. doi: 10.1038/s41598-024-64608-1

13. Zou J, Tran D, Baalbaki M, Tang LF, Poon A, Pelonero A, et al. An internal promoter underlies the difference in disease severity between N- and C-terminal truncation mutations of Titin in zebrafish. Elife 2015;4:e09406. doi: 10.7554/eLife.09406

14. Golbus JR, Puckelwartz MJ, Fahrenbach JP, Dellefave-Castillo LM, Wolfgeher D, McNally EM. Population-based variation in cardiomyopathy genes. Circ Cardiovasc Genet 2012;5:391–399. doi: 10.1161/CIRCGENETICS.112.962928

15. Akinrinade O, Koskenvuo JW, Alastalo TP. Prevalence of Titin Truncating Variants in General Population. PLoS One 2015;10:e0145284. doi: 10.1371/journal.pone.0145284

16. Herman DS, Lam L, Taylor MR, Wang L, Teekakirikul P, Christodoulou D, et al. Truncations of titin causing dilated cardiomyopathy. N Engl J Med 2012;366:619–628. doi: 10.1056/NEJMoa1110186

17. Morales A, Kinnamon DD, Jordan E, Platt J, Vatta M, Dorschner MO, et al. Variant Interpretation for Dilated Cardiomyopathy: Refinement of the American College of Medical Genetics and Genomics/ClinGen Guidelines for the DCM Precision Medicine Study. Circ Genom Precis Med 2020;13:e002480. doi: 10.1161/CIRCGEN.119.002480

18. Vatta M, Regalado E, Parfenov M, Swartzlander D, Nagl A, Mannello M, et al. Analysis of TTN Truncating Variants in >74 000 Cases Reveals New Clinically Relevant Gene Regions. Circ Genom Precis Med 2025;18:e004982. doi: 10.1161/CIRCGEN.124.004982

19. Shah RA, Asatryan B, Sharaf Dabbagh G, Aung N, Khanji MY, Lopes LR, et al. Frequency, Penetrance, and Variable Expressivity of Dilated Cardiomyopathy-Associated Putative Pathogenic Gene Variants in UK Biobank Participants. Circulation 2022;146:110–124. doi: 10.1161/CIRCULATIONAHA.121.058143

20. Massier M, de Groote P, Donal E, Magnin-Poull I, Coubes C, Le Guillou Horn X, et al. Exploring the Familial Phenotypic Variability Associated With TTN Truncating Variants in Cardiomyopathies: Variant Spectrum, Genotype-Phenotype Correlation and Consequences in Genetic Counseling. Clin Genet 2025;107:425–433. doi: 10.1111/cge.14679

21. Jani VP, Yoo EJ, Binek A, Guo A, Kim JS, Aguilan J, et al. Myocardial Proteome in Human Heart Failure With Preserved Ejection Fraction. J Am Heart Assoc 2025;14:e038945. doi: 10.1161/JAHA.124.038945

22. Cao L, Rushakoff J, Williamson I, Karlstaedt A, Kittleson M, Czer L, et al. Similar burden of rare genetic variants in ischemic and non-ischemic dilated cardiomyopathy. Front Cardiovasc Med 2025;12:1542653. doi: 10.3389/fcvm.2025.1542653

23. Jordan E, Peterson L, Ai T, Asatryan B, Bronicki L, Brown E, et al. Evidence-Based Assessment of Genes in Dilated Cardiomyopathy. Circulation 2021;144:7–19. doi: 10.1161/CIRCULATIONAHA.120.053033

24. McLaren W, Gil L, Hunt SE, Riat HS, Ritchie GR, Thormann A, et al. The Ensembl Variant Effect Predictor. Genome Biol 2016;17:122. doi: 10.1186/s13059-016-0974-4

25. Franklin by Qiagen. https://franklin.genoox.com

26. Deep 6 AI. https://deep6.ai/

27. Gallone G, Veen KM, Bernhardt AM, Caliskan K, Najjar E, Netuka I, et al. Long-Term Outcomes of Contemporary Durable Left Ventricular Assist Device Support in Younger Patients: A EUROMACS Analysis. Eur J Cardiothorac Surg 2026;68. doi: 10.1093/ejcts/ezag234

28. Truby LK, Farr MA, Garan AR, Givens R, Restaino SW, Latif F, et al. Impact of Bridge to Transplantation With Continuous-Flow Left Ventricular Assist Devices on Posttransplantation Mortality. Circulation 2019;140:459–469. doi: 10.1161/CIRCULATIONAHA.118.036932

29. Richards D, Cole RM, Kittleson M, Mitchell D, Yazdi V, Chonde M, et al. Enhanced structural reverse remodeling following LVAD support in patients with TTN truncating variants. JHLT Open 2026;14:100688. doi: 10.1016/j.jhlto.2026.100688

30. Wilcox JE, Fang JC, Margulies KB, Mann DL. Heart Failure With Recovered Left Ventricular Ejection Fraction: JACC Scientific Expert Panel. J Am Coll Cardiol 2020;76:719–734. doi: 10.1016/j.jacc.2020.05.075

31. Kinnamon DD, Morales A, Bowen DJ, Burke W, Hershberger RE, Consortium* DCM. Toward Genetics-Driven Early Intervention in Dilated Cardiomyopathy: Design and Implementation of the DCM Precision Medicine Study. Circ Cardiovasc Genet 2017;10. doi: 10.1161/CIRCGENETICS.117.001826

32. Li H. Minimap2: pairwise alignment for nucleotide sequences. Bioinformatics 2018;34:3094–3100. doi: 10.1093/bioinformatics/bty191

33. Chen Y, Sim A, Wan YK, Yeo K, Lee JJX, Ling MH, et al. Context-aware transcript quantification from long-read RNA-seq data with Bambu. Nat Methods 2023;20:1187–1195. doi: 10.1038/s41592-023-01908-w

34. Wissel D, Mehlferber MM, Nguyen KM, Pavelko V, Tseng E, Robinson MD, et al. A Systematic Benchmark of High-Accuracy PacBio Long-Read RNA Sequencing for Transcript-Level Quantification. bioRxiv 2025. doi: 10.1101/2025.05.30.656561

35. Zare Jousheghani Z, Singh NP, Patro R. Oarfish: enhanced probabilistic modeling leads to improved accuracy in long read transcriptome quantification. Bioinformatics 2025;41:i304–i313. doi: 10.1093/bioinformatics/btaf240

36. Love MI, Huber W, Anders S. Moderated estimation of fold change and dispersion for RNA-seq data with DESeq2. Genome Biol 2014;15:550. doi: 10.1186/s13059-014-0550-8

37. Dobin A, Davis CA, Schlesinger F, Drenkow J, Zaleski C, Jha S, et al. STAR: ultrafast universal RNA-seq aligner. Bioinformatics 2013;29:15–21. doi: 10.1093/bioinformatics/bts635

38. Huber W, Carey VJ, Gentleman R, Anders S, Carlson M, Carvalho BS, et al. Orchestrating high-throughput genomic analysis with Bioconductor. Nat Methods 2015;12:115–121. doi: 10.1038/nmeth.3252

39. H P. Biostrings.

40. Wang ET, Sandberg R, Luo S, Khrebtukova I, Zhang L, Mayr C, et al. Alternative isoform regulation in human tissue transcriptomes. Nature 2008;456:470–476. doi: 10.1038/nature07509

41. Demichev V, Messner CB, Vernardis SI, Lilley KS, Ralser M. DIA-NN: neural networks and interference correction enable deep proteome coverage in high throughput. Nat Methods 2020;17:41–44. doi: 10.1038/s41592-019-0638-x

42. Computing RFfS.

43. Wickham H. ggplot2: Elegant Graphics for Data Analysis: Springer International Publishing; 2016.

44. Shahid A, Kim J, Byon SS, Hong S, Lee I, Lee BD. An end-to-end pipeline for automated scoliosis diagnosis with standardized clinical reporting using SNOMED CT. Sci Rep 2025;15:17274. doi: 10.1038/s41598-025-01952-w

45. Clayton JA, Collins FS. Policy: NIH to balance sex in cell and animal studies. Nature 2014;509:282–283. doi: 10.1038/509282a

46. Friedman J, Hastie T, Tibshirani R. Regularization Paths for Generalized Linear Models via Coordinate Descent. J Stat Softw 2010;33:1–22.

47. Collins GS, Dhiman P, Ma J, Schlussel MM, Archer L, Van Calster B, et al. Evaluation of clinical prediction models (part 1): from development to external validation. BMJ 2024;384:e074819. doi: 10.1136/bmj-2023-074819

48. Robin X, Turck N, Hainard A, Tiberti N, Lisacek F, Sanchez JC, et al. pROC: an open-source package for R and S+ to analyze and compare ROC curves. BMC Bioinformatics 2011;12:77. doi: 10.1186/1471-2105-12-77

49. Van Calster B, McLernon DJ, van Smeden M, Wynants L, Steyerberg EW, Topic Group ’Evaluating diagnostic t, et al. Calibration: the Achilles heel of predictive analytics. BMC Med 2019;17:230. doi: 10.1186/s12916-019-1466-7

50. Leening MJ, Vedder MM, Witteman JC, Pencina MJ, Steyerberg EW. Net reclassification improvement: computation, interpretation, and controversies: a literature review and clinician’s guide. Ann Intern Med 2014;160:122–131. doi: 10.7326/M13-1522

51. Kundu S, Aulchenko YS, van Duijn CM, Janssens AC. PredictABEL: an R package for the assessment of risk prediction models. Eur J Epidemiol 2011;26:261–264. doi: 10.1007/s10654-011-9567-4

52. Akhtar MM, Lorenzini M, Pavlou M, Ochoa JP, O’Mahony C, Restrepo-Cordoba MA, et al. Association of Left Ventricular Systolic Dysfunction Among Carriers of Truncating Variants in Filamin C With Frequent Ventricular Arrhythmia and End-stage Heart Failure. JAMA Cardiol 2021;6:891–901. doi: 10.1001/jamacardio.2021.1106

53. Haggerty CM, Damrauer SM, Levin MG, Birtwell D, Carey DJ, Golden AM, et al. Genomics-First Evaluation of Heart Disease Associated With Titin-Truncating Variants. Circulation 2019;140:42–54. doi: 10.1161/CIRCULATIONAHA.119.039573

54. Shetty NS, Pampana A, Patel N, Li P, Arora G, Arora P. High-proportion spliced-in titin truncating variants in African and European ancestry in the All of Us Research Program. Nat Cardiovasc Res 2024;3:140–144. doi: 10.1038/s44161-023-00417-5

55. DePaolo J, Bornstein MR, Judy R, Abramowitz S, Verma SS, Levin MG, et al. Titin-Truncating variants predispose to dilated cardiomyopathy in populations genetically similar to african and european reference populations. PLoS Genet 2025;21:e1011727. doi: 10.1371/journal.pgen.1011727

56. Ntusi NAB, Sliwa K. Impact of Racial and Ethnic Disparities on Patients With Dilated Cardiomyopathy: JACC Focus Seminar 7/9. J Am Coll Cardiol 2021;78:2580–2588. doi: 10.1016/j.jacc.2021.10.021

57. Johnson R, Fletcher RA, Peters S, Ohanian M, Soka M, Smolnikov A, et al. Titin-related familial dilated cardiomyopathy: factors associated with disease onset. Eur Heart J 2025. doi: 10.1093/eurheartj/ehaf380

58. Li Y, Xian H, Xu Y, Li W, Guo J, Wan K, et al. The impact of type 2 diabetes mellitus on the clinical profile, myocardial fibrosis, and prognosis in non-ischemic dilated cardiomyopathy: a prospective cohort study. Cardiovasc Diabetol 2024;23:48. doi: 10.1186/s12933-024-02134-0

59. Vikhorev PG, Vikhoreva NN, Yeung W, Li A, Lal S, Dos Remedios CG, et al. Titin-truncating mutations associated with dilated cardiomyopathy alter length-dependent activation and its modulation via phosphorylation. Cardiovasc Res 2022;118:241–253. doi: 10.1093/cvr/cvaa316

60. Helms AS, Tang VT, O’Leary TS, Friedline S, Wauchope M, Arora A, et al. Effects of MYBPC3 loss-of-function mutations preceding hypertrophic cardiomyopathy. JCI Insight 2020;5. doi: 10.1172/jci.insight.133782

