## Supplemental Methods and Figure for "Exon Proteogenomic Abundance Predicts Advanced Heart Failure in Titin Cardiomyopathy"

**Table of Contents**

**Supplemental Methods**

**Supplemental Figure 1**

**Supplemental Methods**

*Quality Assessment of Long-read RNA Sequencing for TTN*

Read coverage (5’ to 3’) was assessed using RSeQC ^1^ and coverage for *TTN* showed no 3’ bias with a median 5’-to-3’ coverage ratio of 1.06 (Supplemental Table 6). Internal priming was assessed with Pigeon (a PacBio-specific implementation of SQUANTI3 ^2^) and results showed that internal priming for *TTN* affected a median of 5% of transcripts (interquartile range 3.9-9.3; Supplemental Table 7). Note that due to the extreme length of *TTN* transcripts compared to the maximum length of PacBio long-read RNA sequencing reads, internal priming is needed to reach the 5’ end of these transcripts. Exon-junction coverage, expressed as a percentage of total *TTN* reads, was assessed using a bespoke R script and showed consistent identification of exon-junctions, with only 8 junctions missing from all samples (Supplemental Table 8). We compared long-read RNA sequencing exon-junction coverage to that of short-read RNA sequencing and found systematic inflation of *TTN* coverage by short-read sequencing (Supplemental Table 8): the median *TTN* exon-junction coverage was higher by short-read RNA sequencing as compared to long-read sequencing for 176/381 (46%) of the exon-junctions of the *TTN* metatranscript, equal for 62/381 (16%), and lower for (23%).

*Spatial Transcriptomics*

Formalin-fixed paraffin-embedded slides from 4 heart explants were obtained: 1 from an organ donor not utilized for HT, 1 from a patient undergoing HT for end-stage DCM due to a *DSP* variant, and 2 patients undergoing HT for end-stage DCM due to a TTN-TV. The Xenium Human Immuno-Oncology Profiling Panel was utilized with the addition of custom probes for the *TTN* isoforms N2BA, Novex-3, and metatranscript (designed to span a unique exon-exon junction), *DSP*, and the housekeeping gene glyceraldehyde 3-phosphate dehydrogenase (*GAPDH*). Custom probe sequences are provided in Supplemental Table 6. Probe hybridization, ligation, and rolling circle amplification were performed, following the manufacturer’s protocol. Imaging and signal decoding were done using the Xenium Analyzer instrument (10x Genomics). Each slide’s Xenium output was exported as a “spatialexperiment” object and normalized using SpaNorm ^3^. Normalized counts for each slide were rescaled to between 0 and 1000 and plotted using the R package Voyager ^4^.

*Mass Spectrometry-based Proteomics*

28 samples underwent proteomic mass spectrometry (MS) sample processing as previously reported ^5^. Briefly, protein was isolated from explanted cardiac tissue by the “in sequence” method ^6^ *i.e.*: each tissue was fractionated into cytosolic-, myofilament-, and insoluble-enriched protein fractions, and trypsin digested on S-Trap columns (ProtiFi). Peptides from each patient sample were analyzed in triplicate by liquid chromatography (LC)-MS/MS in data independent acquisition (DIA) mode on an Ultimate 3000 NanoLC connected to an Orbitrap Fusion™ Lumos™ Tribrid™ Mass Spectrometer (Thermo Fisher Scientific) equipped with an EasySpray ion source. Data acquisition was completed blinded to experimental groups. LC-MS/MS method was adapted and modified from previously published workflow ^7^. Peptides were separated on a C18 column (15 cm with 300 µm ID, 3 µm Omega Polar C18 beads, and 100 Ǻ pore size, Phenomenex) over the course of total 90 minutes at a flow rate of 8µL/min. Mobile phase A was 0.1% v/v formic acid in water and mobile phase B was 90% ACN, 0.1% formic acid in water. Initial loading condition was 1% B for 3 min and then 3% for another 2 min at a flow rate of 8 μL/min to allow for equilibration and for samples to reach the column, followed by a linear gradient of 3–30% B over 70 min, 30–40% B over 10 min and 40–45% B over 5 min. Each MS1 scan was followed by 150 DIA MS2 scans ^8^ (4Da size precursor window).

*DIA-NN*

Peptide identification and quantification was performed using DIA-NN ^9^. The whole proteome sequence (excluding the canonical *TTN* metatranscript) was supplemented with the protein sequences of the 15 human *TTN* isoforms (Supplemental Table 12) and was used to generate the spectral library. *In silico* digestion was performed using cleavage at K and R. A deep-learning model was applied to generate an in silico spectral library from the resulting peptide list. The fragment m/z range was set to 200–1800. Peptide length was restricted to 7–40 amino acids. The precursor m/z range was set to 300–1800, and precursor charge states were limited to 1–4. Cysteine carbamidomethylation was included as a fixed modification, and N-terminal methionine was excluded. A maximum of one missed cleavage was allowed. Quantification used a fixed-width center of each elution peak. DIA-NN automatically optimized mass accuracy using the first run of the experiment. RT-dependent cross-run normalization was applied, and results were filtered at a 0.01 FDR threshold.

*Proteoform Mapping*

The 15 human *TTN* isoforms (Supplemental Table 12) were digested *in silico* with trypsin with allowance of one missed cleavage. Unique peptides, present in only a single isoform, were selected to give a unique peptide map. The unique peptide map was compared to experimentally identified peptides with an intensity above 0 in at least one sample.

*Web Application Development*

The TTN Variant Recoder and Annotator application was developed as an interactive web application using R Shiny. The app combines a reactive client-side user interface with server-side data integration. At runtime, the application queries the Ensembl REST API variant recoder (https://rest.ensembl.org/documentation/info/variant_recoder) and variant effect predictor (https://rest.ensembl.org/documentation/info/vep_hgvs_get) to extract HGVS nomenclature and consequence terms, mapping them to standardized clinical categories (frameshift, stop gain, splice, and other) and resolving exon/intron usage. It dynamically cross-references these annotations with files containing the PSI-Original and QPI-PSI values, as well as the multivariable model odds ratios (hosted on GitHub), to compute the probability of AHF. The application is hosted via the Posit Connect platform.

*References*

1.	Wang L, Wang S, Li W. RSeQC: quality control of RNA-seq experiments. *Bioinformatics* 2012;**28**:2184–2185. doi: <https://doi.org/10.1093/bioinformatics/bts356>

2.	Pardo-Palacios FJ, Arzalluz-Luque A, Kondratova L, Salguero P, Mestre-Tomas J, Amorin R, et al. SQANTI3: curation of long-read transcriptomes for accurate identification of known and novel isoforms. *Nat Methods* 2024;**21**:793–797. doi: <https://doi.org/10.1038/s41592-024-02229-2>

3.	Salim A, Bhuva DD, Chen C, Tan CW, Yang P, Davis MJ, et al. SpaNorm: spatially-aware normalization for spatial transcriptomics data. *Genome Biol* 2025;**26**:109. doi: <https://doi.org/10.1186/s13059-025-03565-y>

4.	Moses L, Einarsson PH, Jackson K, Luebbert L, Booeshaghi AS, Antonsson S, et al. Voyager: exploratory single-cell genomics data analysis with geospatial statistics. *bioRxiv* 2023. doi: <https://doi.org/10.1101/2023.07.20.549945>

5.	Meddeb M, Koleini N, Binek A, Keykhaei M, Darehgazani R, Kwon S, et al. Myocardial ultrastructure of human heart failure with preserved ejection fraction. *Nat Cardiovasc Res* 2024;**3**:907–914. doi: <https://doi.org/10.1038/s44161-024-00516-x>

6.	Kane LA, Neverova I, Van Eyk JE. Subfractionation of heart tissue: the "in sequence" myofilament protein extraction of myocardial tissue. *Methods Mol Biol* 2007;**357**:87–90. doi: <https://doi.org/10.1385/1-59745-214-9:87>

7.	Mc Ardle A, Binek A, Moradian A, Chazarin Orgel B, Rivas A, Washington KE, et al. Standardized Workflow for Precise Mid- and High-Throughput Proteomics of Blood Biofluids. *Clin Chem* 2022;**68**:450–460. doi: <https://doi.org/10.1093/clinchem/hvab202>

8.	Robinson AE, Binek A, Venkatraman V, Searle BC, Holewinski RJ, Rosenberger G, et al. Lysine and Arginine Protein Post-translational Modifications by Enhanced DIA Libraries: Quantification in Murine Liver Disease. *J Proteome Res* 2020;**19**:4163–4178. doi: <https://doi.org/10.1021/acs.jproteome.0c00685>

9.	Demichev V, Messner CB, Vernardis SI, Lilley KS, Ralser M. DIA-NN: neural networks and interference correction enable deep proteome coverage in high throughput. *Nat Methods* 2020;**17**:41–44. doi: <https://doi.org/10.1038/s41592-019-0638-x>

**Supplemental Figure 1**

**A**

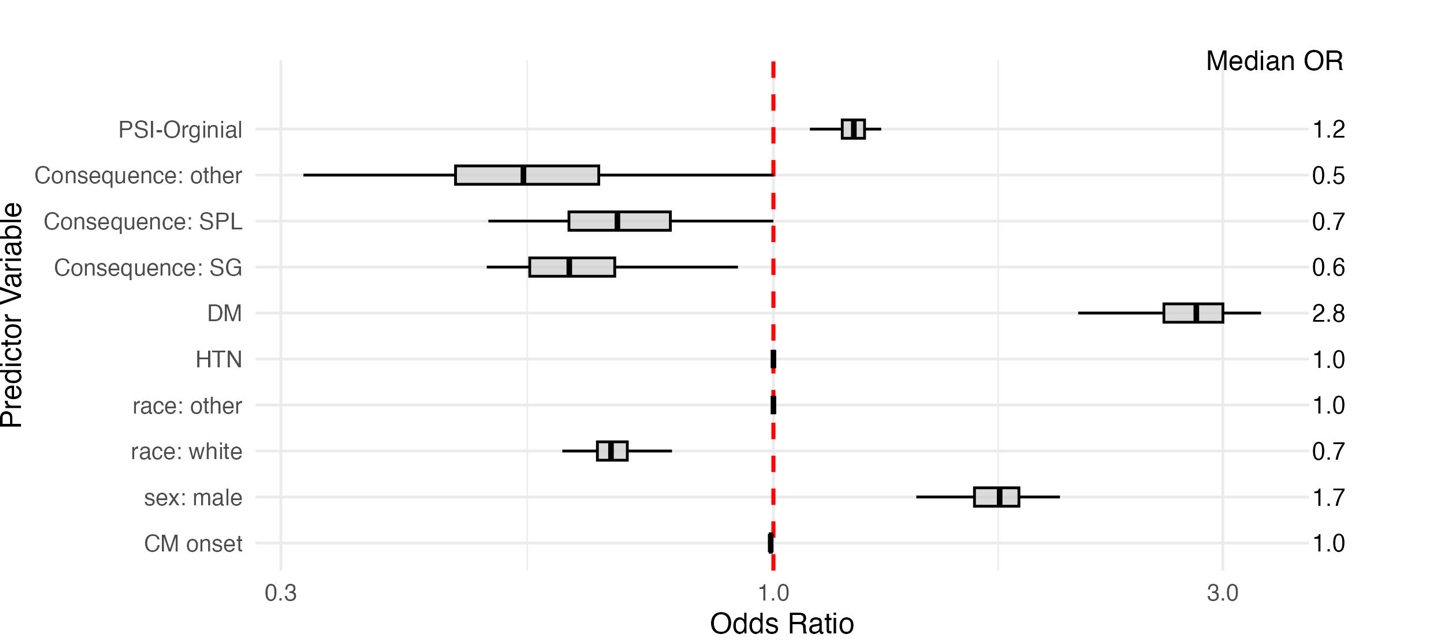

**B**

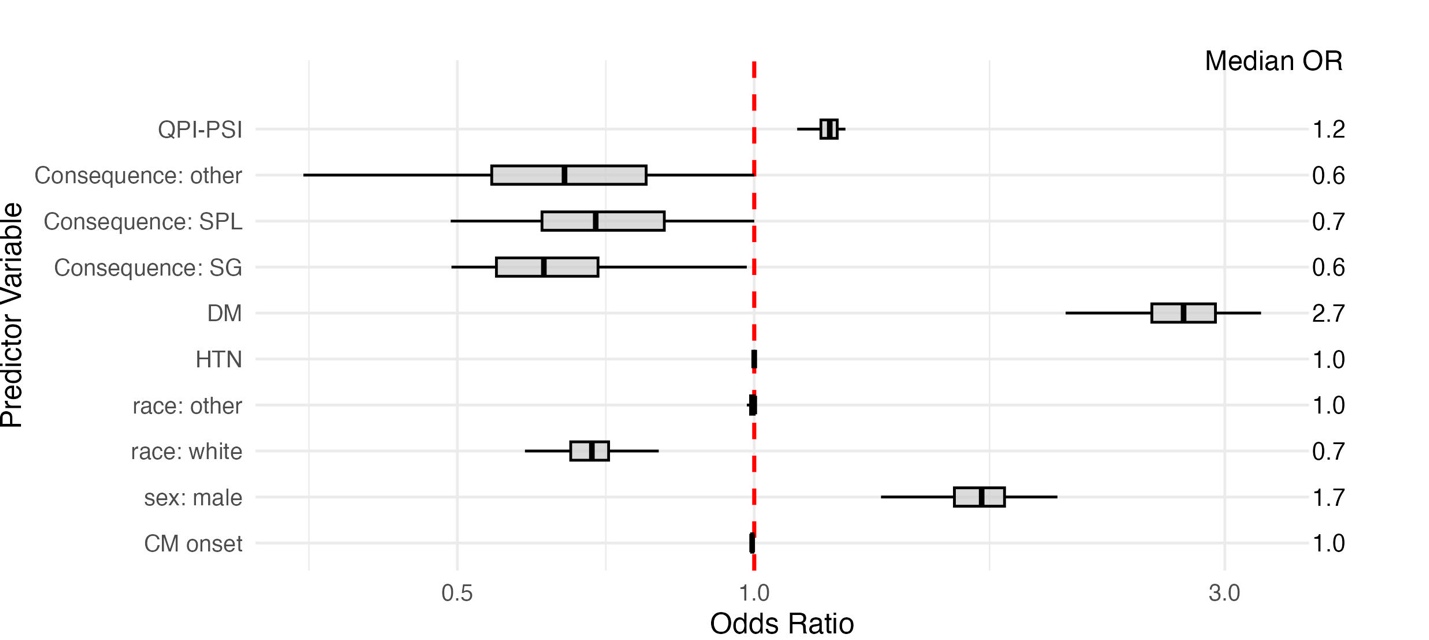

**Figure S1:** (A) Forest plot of odds ratios from a multivariable logistic model for advanced heart failure including PSI-Original. Grey box indicates interquartile range and line inside the box indicates the median. (B) Forest plot of odds ratios from a multivariable logistic model for advanced heart failure including QPI-PSI. Grey box indicates interquartile range and line inside the box indicates the median.
